# The causal relationships between leisure-time physical activity and body mass index in adulthood: A triangulation study

**DOI:** 10.64898/2026.03.10.26348015

**Authors:** Anna Kankaanpää, Laura Joensuu, Ulf Ekelund, Anni Pitkänen, Katja Waller, Teemu Palviainen, Jaakko Kaprio, Miina Ollikainen, Sari Aaltonen, Elina Sillanpää

**Author notes:** Corresponding author: Anna Kankaanpää, Postdoctoral researcher Faculty of Sport and Health Sciences, University of Jyväskylä, P.O. Box 35 (VIV), FIN-40014 Jyväskylä, Finland.

## Abstract

**Background:** Previous studies have presented conflicting findings regarding the potential causal relationships between leisure-time physical activity (LTPA) and body mass index (BMI). Here, we use individual-level data and apply a triangulation framework that incorporates three complementary methods to investigate the bidirectional causal associations between LTPA and BMI.

**Methods:** We used data from a longitudinal Finnish twin cohort with four measurement points spanning 36 years. The data included 22,696 twin individuals aged 18-50 years at baseline (52.4% women); 8,527 had genetic data available. We applied three analytical approaches suggested to strengthen causal inference in observational studies: Random intercept cross-lagged path model (RI-CLPM) for longitudinal data, one-sample Mendelian Randomization (MR) and Direction of Causation (DoC and MR-DoC) twin models for cross-sectional data at each measurement point.

**Results:** All three approaches provided evidence for a causal effect of higher BMI on lower LTPA, particularly at the later follow-up stages. Only twin models suggested a negative causal effect of LTPA on BMI. Men and women showed mainly similar effects.

**Conclusions:** Evidence triangulation across the three methodologies provided support for a causal effect of higher BMI on lower LTPA, whereas the evidence for a reverse effect was less convincing. Our results indicate that the role of high BMI in limiting LTPA becomes more important with advancing age, while also highlighting the importance of accounting for timing when studying the causal effects of LTPA on BMI and vice versa.

## 1 Introduction

Physical activity (PA) requires additional energy expenditure^1^; therefore, PA is considered to improve weight management while reducing the risk of obesity. Simultaneously, excess body weight may limit PA by impairing the capacity for body movement. Energy expenditure from PA is traditionally categorised into domains (i.e., leisure-time, commuting and work),^2^ with leisure-time viewed as one of the true options for managing self-selected energy expenditure.^3^ Most of the observational evidence in adults indicates an association between higher leisure-time physical activity (LTPA) and lower body mass index (BMI) and lower weight gain, as supported by reviews^4,5^ and findings from twin studies.^6,7^ However, these associations are challenged by some studies that suggest no causality for this relationship.^8,9^ Total energy expenditure (TEE) comprises basal metabolic rate, diet-induced energy expenditure, and energy expenditure from PA, and it remains debated whether increases in PA increase TEE in the long-term, or become compensated by reductions in other components.^1,10^ Additionally, lack of temporal order, the potential for unmeasured confounding, and the possibility for reverse causality could limit interpretations of the findings from the previous observational studies assessing the associations between LTPA and BMI.

Establishing causality requires evidence of the temporal sequence of the traits, which could be provided by longitudinal data.^11^ However, the majority of studies examining LTPA and BMI in adult populations have focused on possible associations between the level of PA (or concurrent changes in PA) and future changes in BMI.^6,12–16^ Notably, many longitudinal studies that have used multiple timepoints and advanced statistical models have consistently demonstrated negative associations between parallel changes in LTPA and BMI, whereas LTPA level alone does not appear to predict subsequent changes in BMI.^16–18^ However, the finding of an association between parallel changes does not allow conclusions regarding the temporal ordering of these changes. The few studies that have investigated bidirectional associations between LTPA and BMI^8,17,19^ have mostly concluded that high BMI is more likely to predict subsequent declines in PA than high PA is to predict reduced weight gain.^8,19^

To improve causal interpretation, a longitudinal model is typically adjusted for potential confounders, including demographic characteristics (e.g., sex, age, socioeconomic status) and lifestyle factors such as diet, smoking and alcohol, and these confounders are typically measured at a single time point.^9^ A better model choice for analysing within-person dynamics is the random intercept cross-lagged panel model (RI-CLPM), as this tool would enable investigation of whether deviations in LTPA from an individual’s own level predict subsequent deviations in BMI, and vice versa.^20^ By separating between- and within-individual level variation, the RI-CLPM accounts for time-invariant confounding, thereby yielding results that are probably less biased by residual and unmeasured confounding, although time-varying confounders may still affect the estimates.

Current data indicate that LTPA and BMI are influenced by heritability (as demonstrated by twin studies) ^21–24^ and genetics (as shown using genome-wide association studies [GWASs]).^25,26^ Therefore, shared genetics could underly both traits, with their association explained by genetic confounding, with individuals with a genetic predisposition to high LTPA more likely to be active and carry genes that help maintain an ideal weight. However, the findings from previous twin studies suggest that the negative genetic correlation between LTPA and BMI is relatively weak (-0.20 to -0.08),^27^ with further support provided by results from measured genetics studies.^28^ The twin design strengthens causal inference because it enables the control of familial factors, including genetics and shared environment, which are rarely accounted for in observational studies.^29^ Previous twin studies have indicated that the association of long-term LTPA with lesser weight gain is not fully accounted for by familial factors.^6,7,27,30^ However, these studies did not examine in detail the potential for effects in the opposite direction. To test directly competing causal hypotheses, the Direction of Causation (DoC) model could be an option. The DoC model can use cross-sectional twin data to establish causality by utilising cross-twin cross-trait correlations.^31^

Another method for testing causal association is Mendelian randomisation (MR), which uses genetic instruments to probe the associations found in observational studies between an exposure and an outcome.^32^ MR studies are described as naturally occurring randomised trials in which genetic factors are randomly assigned by nature. Therefore, confounding and reverse causation are less likely to affect the results obtained using MR than those obtained using traditional observational methods. Indeed, the use of MR is increasingly being reported in the literature on causal links between both self-reported and device-based PA and BMI in adults.^33–38^ However, the findings of these MR studies are mixed, with results ranging from bidirectional negative causal associations^33,35,37,38^ to null findings.^34,36^ Almost all of these studies have used two-sample MR, which utilises summary-level data from GWASs to avoid weak instrument bias. Relying on existing summary-level datasets typically provides overall lifetime estimates for causal effects, but it limits the possibility of examining sex- and time-specific effects.

One issue with the use of MR is its use of individual genetic variant(s) (specifically single nucleotide polymorphisms [SNPs]) as instrumental variables for the phenotypes. Both LTPA and BMI are complex phenotypes that are regulated by multiple genes, and individual genetic variants are only weakly associated with them.^28,39^ Therefore, instead of using individual SNPs as instrumental variables for these phenotypes in MR, it may be more useful to employ polygenic scores (PGS) that sum the effect of multiple SNPs into a single value. However, using PGS as instrument may violate the MR assumption of no horizontal pleiotropy. This limitation could be addressed by using MR-DoC models, which integrate MR with the DoC twin model, allowing polygenic scores to serve as stronger instrumental variables even when they exhibit pleiotropic effects.^40,41^

While progress has been made in disentangling the nature of the LTPA-BMI associations, further advancement requires the adoption of novel methodological approaches. One such approach is evidence triangulation, which helps to address potential biases by integrating findings from multiple study designs and methods that have different underlying assumptions.^42,43^ In the present study, we apply the following triangulation approach to examine the LTPA-BMI associations: First, we use longitudinal data with four measurements and fit an RI-CLPM to assess bidirectional predictive associations. Second, we use cross-sectional LTPA and BMI data to conduct a one-sample MR, employing both PGSs and included SNPs as instrumental variables. Third, we utilise a twin study design to investigate causal effects by applying both traditional DoC models and MR-DoC models, with the latter incorporating PGSs into the model. The MR and twin models are estimated separately at four distinct time points to examine potential time-specific effects. All analyses are additionally stratified by sex to examine sex-specific causal effects.

## 2 Material and Methods

### 2.1 Study design and participants

The older Finnish Twin Cohort (FTC) study was established 50 years ago, and the data collected have been extensively reviewed.^44^ Originally, the FTC consisted of same-sex twin pairs born in Finland before 1958, where both cotwins were alive in 1967. Questionnaires were mailed in 1975 and 1981 to all twins born before 1958 and living in Finland, with a follow-up questionnaire conducted in 1990 with twins born between 1930 and 1957. The fourth wave of data collection was conducted in 2011 with twins born between 1945 and 1957. The response rates were high at each time point (89%, 84%, 77% and 72% respectively). The present study included 22,696 twin individuals aged 18-50 years at baseline who had information on LTPA and BMI available in at least one wave (52.4% women). Of these, 8,527 provided genetic information.

### 2.2 Ethics approval

The data collection and all studies were conducted in accordance with the Declaration of Helsinki. The FTC data collection was approved by the ethics committees of the University of Helsinki (113/E3/01 and 346/E0/05) and Helsinki University Central Hospital (270/13/03/01/2008 and 154/13/03/00/2011).

### 2.3 Measurements

#### 2.3.1 Genotyping, quality control, and imputation

Procedures for genotyping, quality control and imputation were previously described.^45^ Variants with a minor allele frequency greater than 1% and imputation quality (INFO score) better than 0.8 were considered in the present study.

#### 2.3.2 Polygenic scores (PGSs)

The SNPs and their corresponding weights were obtained from the original GWASs for BMI^39^ and LTPA ^26,28,35,46^ (Supplementary Table 1). We used the same significance levels as in the original GWASs for selecting SNPs (in most studies, *p* < 5 × 10−8). The PGSs were calculated for the FTC participants using the score function of plink2, and the PGSs were standardised to mean of zero and variance of unity before further analysis.

For **PGS BMI**, we obtained SNPs and corresponding weights from the GWAS provided by Yengo et al.;^39^ this GWAS has been widely used in previous MR studies.^47^ We used 936 SNPs in our calculations of PGS BMI. Because earlier MR studies relied on diverse GWASs to instrument PA, we explored several candidate **PGSs for LTPA** (Supplementary Table 1).^26,28,35,46^

#### 2.3.3 Assessment of phenotypes

**Body mass index (BMI, kg/m^2^)** was based on self-reported height and weight in 1975, 1981, 1990 and 2011. BMI based on self-reports has been shown to agree well with BMI based on measured values.^48^

**Leisure-time physical activity (LTPA)** was assessed in metabolic equivalent (MET) hours per day (h/day) using a structured, validated questionnaire in 1975, 1981, 1990 and 2011.^6,49,50^ In 1975, 1981 and 2011, the questions concerned the monthly frequency of LTPA and the mean session duration and intensity. The MET index was calculated as the product of the frequency, intensity and duration of leisure activities as well as commuting activities, with the resulting values added together.^6,49^ In 1990, the questionnaire differed slightly, as the participants reported their time spent on LTPA (including commuting PA) at different intensity levels. The MET index was calculated by multiplying the time spent on LTPA by the estimated MET value of each intensity level and adding up the values.^50^ Detailed information on scoring was recently described.^51^

The **zygosity** of the twin pairs was determined using a validated questionnaire-based algorithm,^52^ and was confirmed for a subsample using DNA extracted from blood or saliva samples with a high degree of agreement.^44^

### 2.4 Statistical analysis

The MR analysis was conducted using R (version 4.4.1), and longitudinal models as well as twin models were fitted using Mplus (version 8.2).^53^ When longitudinal data were modelled using a structural equation modelling (SEM) framework, missing data were assumed to be missing at random (MAR), and the full information maximum likelihood estimation method with robust standard errors (MLR) was used. For twin modelling, twins were randomised as twin 1 and twin 2, and complete twin pairs were included (10,275 pairs). For the main MR analysis, twin 1 was selected from each pair that had genetic data, with the data from twin 2 used for replication.

#### 2.4.1 Random-intercept cross-lagged panel model for longitudinal data

RI-CLPM was used to examine within-person reciprocal associations between LTPA and BMI in the full cohort. The reciprocal relations between LTPA and BMI were modelled using within-person deviation terms, which capture temporal fluctuations around an individual’s own mean.^20^ Therefore, in contrast to traditional CLPM, which conflates between-person and within-person processes, RI-CLPM strengthens the ability to draw causal inference from estimated cross-lagged paths.^20^ Standard errors were corrected for dependencies due to family structure. To overcome convergence issues, the variables were standardised before analysis.

#### 2.4.2 Mendelian Randomization (MR)

MR involves strong assumptions, including ‘absence of horizontal pleiotropy’ and ‘relevance’.^32^ Horizontal pleiotropy occurs when the SNPs, used as instrumental variables, influence the outcome independent of the exposure. Meeting the relevance assumption requires that the SNPs used as an instrument are adequately associated with the exposure. Additionally, the instrumental variable should be independent of factors that potentially confound the association of interest. In other words, the instrumental variable and outcome should have no common causes (‘independence’ or ‘exchangeability’ assumption).

To select the instrumental variables for MR, we regressed the observed LTPA/BMI on each candidate PGS at each measurement point and assessed the strength of the instrument using the F statistic. F values less than 10 were considered to indicate weak instrument strength.^54^ First, we conducted instrumental variable regression (IVR) using PGSs as instruments, employing the 2-stage least squares estimation method (2SLS). The model was fitted using the AER package.^55^ We then derived summarised associations from the individual-level data; that is, we calculated the associations of each SNP included in the PGS with both the exposure and the outcome using linear regression. Next, we used several methods to estimate causal effects from the summarised data using the MendelianRandomization R package.^56^ Weighted median, the Inverse-Variance-Weighted (IVW) method,^57^ and MR-Egger were utilised.^58^ These methods are more robust than IVR to violations of MR assumptions. For example, MR-Egger adjusts causal estimates for horizontal pleiotropy.^58^

#### 2.4.3 Twin modelling (DoC and MR-DoC models)

The twin model enables the decomposition of variation in a trait into additive genetic (A), dominant genetic (D), shared environmental (C) and non-shared environmental components (E).^59^ Previous twin studies using the same dataset have not found consistent evidence for shared environmental influences on BMI^60–62^ or LTPA^24^ in adulthood and have concluded that the model that include only A and E components (the AE model) fits the data adequately. Therefore, the AE model for both traits was used in further modelling. A bivariate Cholesky model^59^ was used to estimate the genetic (*r_A_*) and environmental correlations (*r_E_*) between LTPA and BMI.

We applied a series of DoC twin models (see the simplified path diagrams for one twin only in Figure 1 and the path diagrams for both twins in Supplementary Figure 1).^31^ First, we estimated a bidirectional DoC model, which included causal paths from BMI to LTPA (*g_1_*) and from LTPA to BMI (*g_2_*) (Model 1). The paths *g_1_* and *g_2_* were set to zero one at a time and unidirectional DoC models were fitted. The model fit was compared with that of the bidirectional model using the Satorra-Bentler corrected chi-squared difference test. Second, we used a unidirectional model adjusted for between-trait genetic correlation (Model 2). To obtain an identifiable model, the environmental correlation was constrained to zero. Third, we specified MR-DoC models by incorporating PGSs in the unidirectional model.^40^ We ran the model without and with the direct path from PGS to the outcome (Model 3 and Model 4, respectively) to assess potential bias due to horizontal pleiotropy.

**Figure 1.**
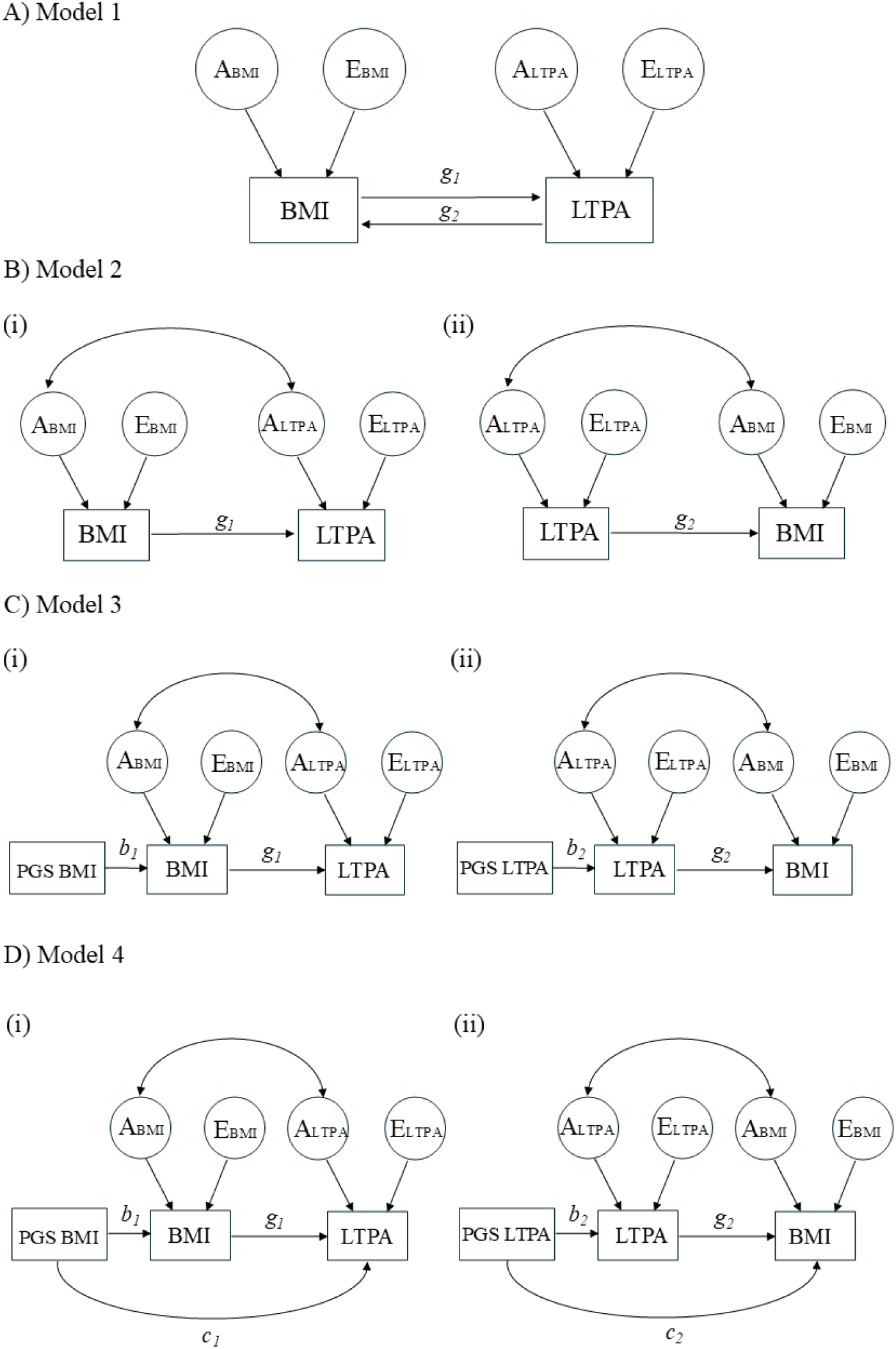
Simplified path diagrams of the direction of causation (DoC) and Mendelian Randomisation (MR)-DoC twin models: A) Bidirectional DoC model, B) Unidirectional DoC model adjusted for genetic correlation, C) MR-DoC model, pleiotropic path constrained to zero, and D) MR-DoC model, pleiotropic path *c* estimated. *Note*. Double-headed arrows represent between-trait genetic correlations. The path diagrams are presented for one twin only.

The twin models at the different measurement points and in three subgroups of twin pairs (all, men and women) were estimated, and the results were extracted using the MplusAutomation package.^63^ To verify the results of MR-DoC models, we used the codes provided by the authors of the original publications^40,41^ (https://cran.r-project.org/web/packages/OpenMx/vignettes/mr.html) and using OpenMx package (version 2.21.11).

Within the SEM framework, we used several model fit indices to evaluate the goodness of fit between the hypothesised model and the observed data. We used the Satorra-Bentler scaled chi-squared test, the comparative fit index (CFI), the Tucker–Lewis index (TLI), the root mean square error of approximation (RMSEA), and the standardised root-mean-square residual (SRMR). The model fits the data well if the chi-squared test is not statistically significant (P > 0.05), the CFI and TLI values are close to 0.95, the RMSEA value is below 0.06, and the SRMR value is below 0.08.^64^

## 3 Results

### 3.1 Descriptive statistics

Table 2 shows the descriptive statistics of the study variables for both the full cohort (with at least one measurement of BMI and LTPA) and the subsample with genetic data.

**Table 1.** Descriptive statistics of the study variables in the full cohort and subsample of twin individuals who had measured genetic data available.

|  | Full cohort (n=22,696) |  | Subsample (n=8,527) |  |
| --- | --- | --- | --- | --- |
|  | n | Mean (SD) or % | n | Mean (SD) or % |
| Zygosity | 22,696 |  | 8,527 |  |
| Unsure | 1,879 | 8.3 | 0 | 0.0 |
| Monozygotic | 6,449 | 28.4 | 2,890 | 33.9 |
| Dizygotic | 14,368 | 63.3 | 5,629 | 66.1 |
| Sex | 22,696 |  | 8,527 |  |
| Women | 11,414 | 50.3 | 4,465 | 52.4 |
| Men | 11,282 | 49.7 | 4,062 | 47.6 |
| Age |  |  |  |  |
| in 1975 | 22,696 | 30.3 (8.9) | 8,527 | 33.0 (8.5) |
| in 1981 | 20,462 | 36.5 (9.0) | 8,092 | 39.1 (8.6) |
| in 1990 | 12,493 | 44.4 (7.8) | 6,188 | 47.3 (7.8) |
| in 2011 | 8,332 | 60.3 (3.8) | 2,801 | 60.5 (3.7) |
| Body mass index (kg/m<sup>2</sup>) |  |  |  |  |
| in 1975 | 20,843 | 22.8 (3.2) | 8,195 | 23.2 (3.2) |
| in 1981 | 20,074 | 23.5 (3.3) | 7,995 | 23.9 (3.4) |
| in 1990 | 12,389 | 24.6 (3.7) | 6,140 | 25.0 (3.8) |
| in 2011 | 8,164 | 26.2 (4.3) | 2,740 | 26.5 (4.4) |
| Leisure-time physical activity |  |  |  |  |
| Metabolic equivalent (MET) index (MET h/day) |  |  |  |  |
| in 1975 | 20,945 | 2.4 (3.1) | 8,231 | 2.3 (2.8) |
| in 1981 | 20,146 | 2.6 (3.1) | 8,016 | 2.5 (3.0) |
| in 1990 | 12,310 | 3.3 (3.4) | 6,107 | 3.1 (3.2) |
| in 2011 | 7,589 | 3.0 (2.8) | 2,563 | 2.8 (2.6) |

SD, standard deviation

Sex-stratified statistics are provided in Supplementary Table 2. In the subsample of twins with measured genetic data, the proportions of monozygotic twins and women were higher, and BMI was higher while LTPA was lower throughout the follow-up compared with the full cohort.

### 3.2 Random-intercept cross-lagged panel model (RI-CLPM)

Figure 2 presents the estimation results of the RI-CLPM based on longitudinal individual-level data on BMI and LTPA. The model fit was acceptable (χ^2^(9)=50.8, RMSEA=0.014; CFI=0.998; TLI=0.995; SRMR=0.008). At the between-person level, the participants reporting higher LTPA over time also reported lower BMI. At the within-person level, the cross-lagged effects indicated that a higher BMI in 1981 was associated with a lower LTPA in 1990, and a higher BMI in 1990 was associated with a lower LTPA in 2011. A higher LTPA in 1975 was associated with a higher BMI in 1981, but no other cross-lagged effects were observed. At the within-person level, the negative residual correlation between BMI and LTPA appeared to become more relevant with age.

**Figure 2.**
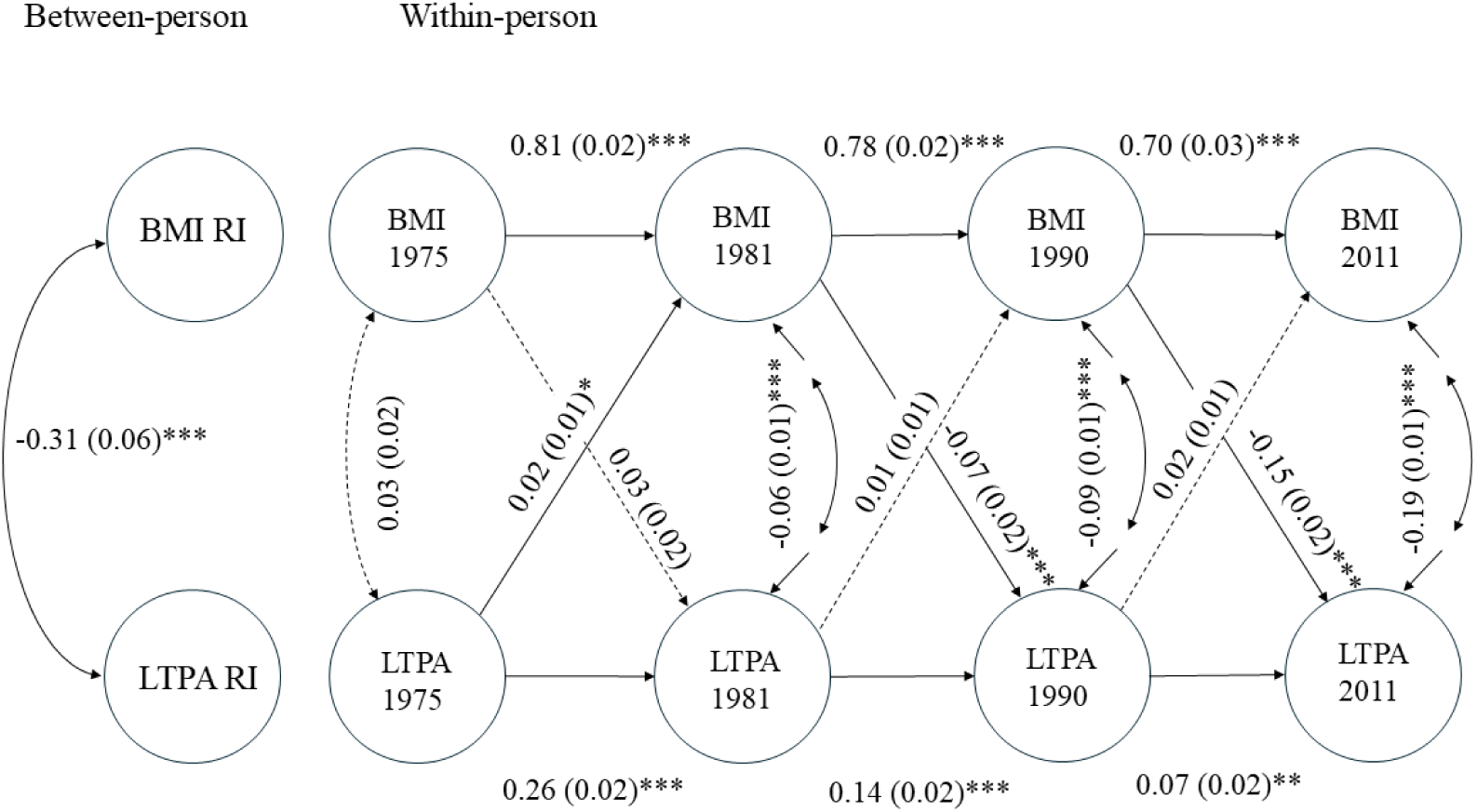
Estimation results of the random-intercept cross-lagged panel models (RI-CLPMs) in full cohort (N = 22,696). The circles denote latent variables and the dashed lines denote associations with no statistical significance (*p* > 0.05). Standardised estimates (standard errors) are presented. \*\*\**p* < 0.001, \*\**p* < 0.01, \**p* < 0.05.

When the analysis was stratified by sex (Supplementary Figure 2), a higher BMI at the within-person level in 1981 was associated with a lower LTPA in 1990 in both men and women, whereas a higher BMI in 1990 was associated with a lower LTPA in 2011 only in women. A higher LTPA in 1990 was associated with a higher BMI in 2011 in men, but no other cross-lagged effects were observed.

### 3.3 Mendelian Randomisation

Pearson’s correlations between the study variables for participants with available genetic data are presented in a heatmap in Supplementary Figure 3, with sex-stratified results shown in Supplementary Figure 4. Restricting the sample to twin 1 only within a twin pair that had genetic data for MR, resulted in the sample consisting of 3,655 individuals. The PGS BMI predicted the observed BMI well at each measurement point. *F* statistics were well above 10 (Supplementary Figure 5). In contrast, the *F* statistics were low (<8) for each PGS considered to represent a candidate for an instrumental variable for LTPA (Supplementary Figures 6, 7 and 8 for all individuals, men and women, respectively). Overall, their associations with the self-reported LTPA in 1975, 1981, 1990 and 2011 were inconsistent and varied around zero, depending on the measurement point and sex. Replication using data from twin 2 yielded results that were as poor as those from the main analysis (Supplementary Figure 9).

Eventually, the PGS for self-reported dichotomised moderate-to-vigorous physical activity (PGS MVPA)^28^ was chosen for further analyses, because it is based on the meta-analysis of GWASs with the largest sample size to date and was used in a recent two-sample MR study.^37^ We also calculated the corresponding genome-wide PGS (see Herranen et al.^65^ for a detailed description of the calculation pipeline [SBayesR]), but we observed no consistent benefit in the instrument’s strength (Supplementary Figures 6-9).

For all participants, the use of PGS BMI as the instrument in IVR indicated a causal effect of higher BMI on lower LTPA but only at the latest measurement point (Figure 3 A). The causal estimates from the Weighted median, IVW and MR-Egger methods were obtained using summarised statistics. These estimates suggested a consistent negative causal effect of BMI on LTPA (i.e. higher BMI led to low LTPA), which appeared to strengthen over time (see also Supplementary Figures 10 A-D).

**Figure 3.**
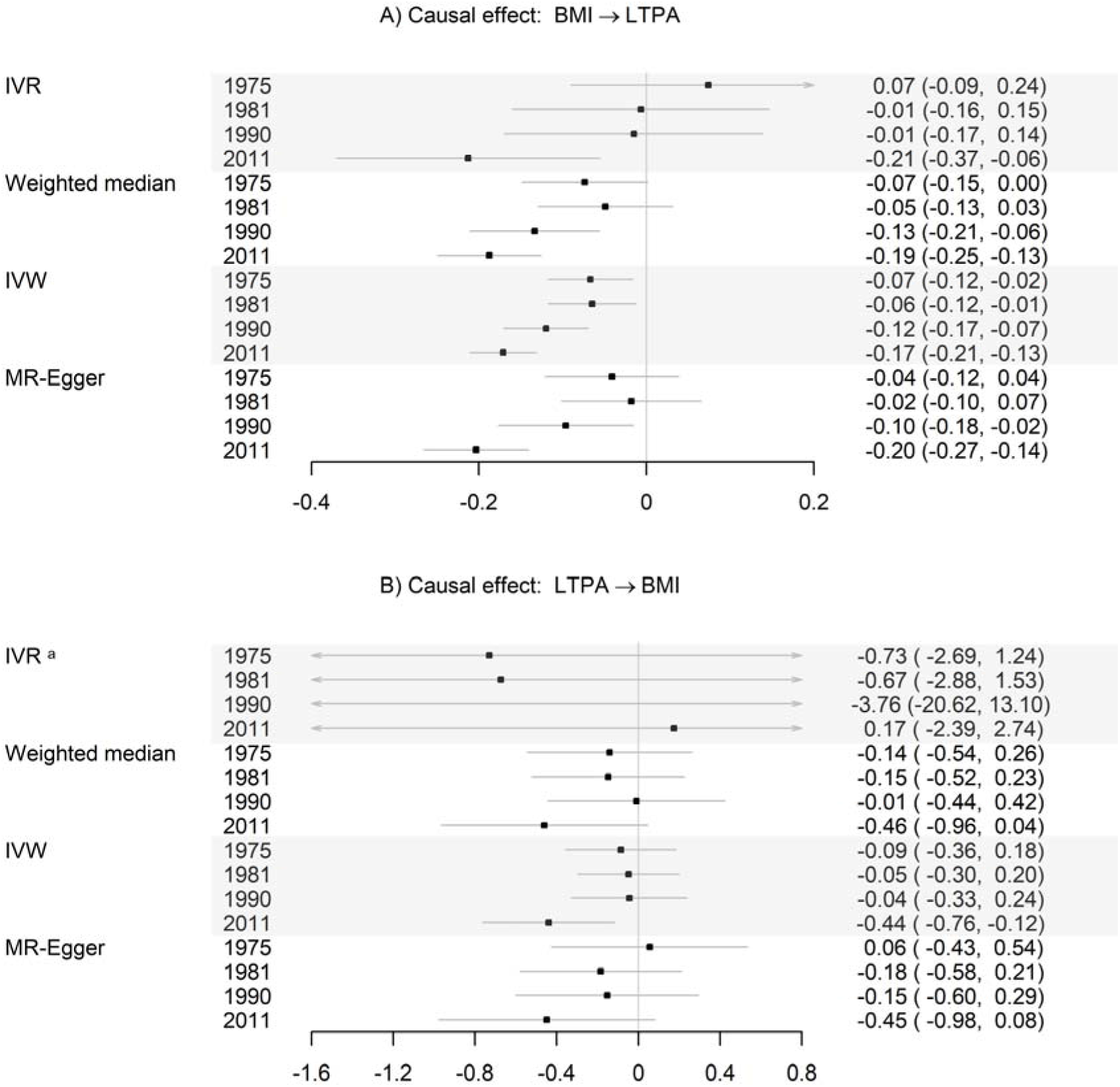
Causal effects (95% confidence intervals) of A) body mass index (BMI) on leisure-time physical activity (LTPA) and B) LTPA on BMI, estimated using several Mendelian randomisation (MR) methods. IVR, Instrumental Variable Regression; IVW, Inverse Variance Weighted. Unstandardised coefficients are presented. Sample sizes at the four time points (1975, 1981, 1990 and 2011) were 3,517; 3,416; 2,715 and 1,164, respectively. Note. ^a^ PGS MVPA was a poor instrument for LTPA.

When PGS MVPA was used as an instrument in IVR, the estimation yielded extremely wide confidence intervals, likely due to weak instrument strength (Figure 3 B). Based on the estimates obtained using summarised statistics, the causal estimate from higher LTPA on lower BMI was consistently negative in 2011, but it did not consistently reach statistical significance across the methods used (see Supplementary Figure 10 E-H).

The interpretation of the results was very similar when the analysis was replicated using data from twin 2 (Supplementary Figure 11). However, when the summarised statistics were used, the causal estimates from LTPA on BMI were consistently negative in 1990 and 2011 and also reached statistical significance (Supplementary Figure 11 B). When the analysis was stratified by sex, a consistent negative causal effect of BMI on LTPA was observed for both men and women, but it strengthened over time only in women (Supplementary Figures 12-15). We found no clear evidence for a causal effect from LTPA to BMI in either men or women. MR-Egger indicated confounding due to the pleiotropic effects between LTPA and BMI in 2011 for both sexes (Supplementary Figures 12 B, 13 B and 15 D, H).

### 3.4 Twin modelling

The phenotypic negative correlation as well as the genetic and unique environmental correlation, between BMI and LTPA appeared to strengthen with time (Supplementary Table 3). The model fits of the DoC models are presented in the Supplementary Table 4. For all twin pairs, the chi-squared difference test indicated a poorer fit in 1981 when the path from LTPA to BMI was constrained to zero (*p* = 0.003), suggesting a causal effect of LTPA on BMI in that year. In contrast, fixing the causal path from BMI to LTPA to zero in 1990 and 2011 resulted in a significantly poorer model fit (*p* < 0.001 in both years). However, the model-fit was acceptable for all models; therefore, we proceeded to fit subsequent models in both directions.

The bidirectional DoC model suggested a causal effect of higher BMI on lower LTPA in 1975, 1990 and 2011 (Figure 4 A, Model 1). Based on the unidirectional DoC models adjusted for between-trait genetic correlation, we observed rather consistent negative causal effects of BMI on LTPA, except in year 1981 (Model 2). Controlling for horizontal pleiotropy in the MR-DoC model resulted in an apparent strengthening of the negative causal path from BMI to LTPA in 1981 (Model 4).

**Figure 4.**
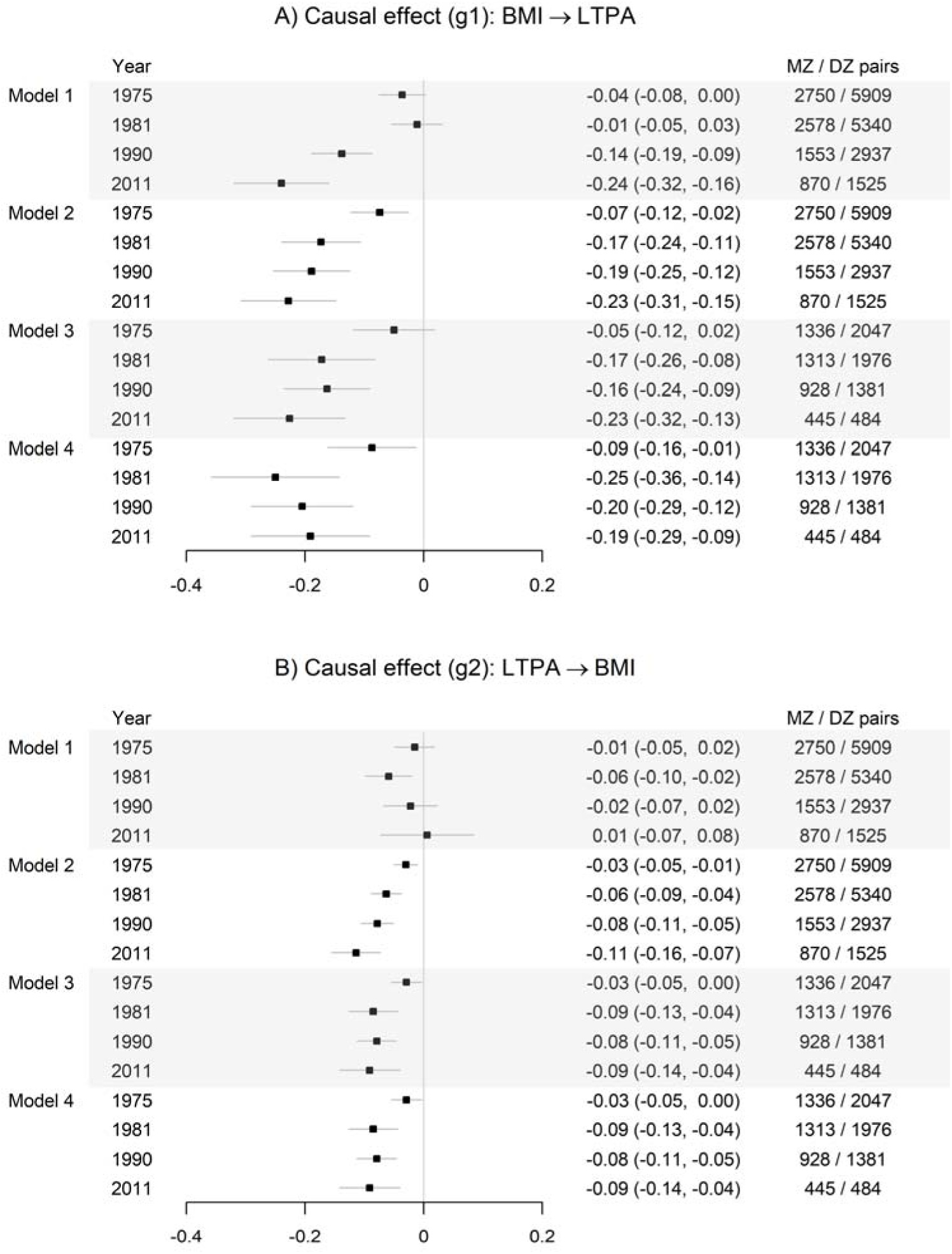
Causal effects of A) body mass index (BMI) on leisure-time physical activity (LTPA) and B) LTPA on BMI in the years 1975, 1981, 1990 and 2011 assessed using Direction of Causation (DoC) and Mendelian Randomization (MR)-DoC twin models. Standardised coefficients (95% confidence intervals) are presented. MZ, monozygotic; DZ, dizygotic. Model 1, bidirectional DoC twin model. Model 2, unidirectional DoC model adjusted for genetic correlation. Model 3, MR-DoC model, no horizontal pleiotropy assumed. Model 4, MR-DoC model, horizontal pleiotropy estimated.

The bidirectional DoC model suggested a causal effect of LTPA on BMI only in 1981 (Figure 4 B, Model 1). The unidirectional DoC and MR-DoC models indicated that higher LTPA was causally associated with lower BMI in 1981, 1990 and 2011 (Models 2-4), but with substantially smaller effect sizes than those observed in the opposite direction. Including a weak instrument and its pleiotropic path in the models did not affect the results (Models 3-4).

After stratifying the analysis by sex, constraining the causal path from BMI to LTPA to zero worsened the model fit in both men and women in 1975 (*p* = 0.001 and *p* = 0.008, respectively), as well as in 1990 and 2011 (all *p* < 0.001). In women, the chi-squared difference test indicated poorer fit in 1981 when the path from LTPA to BMI was constrained to zero (*p* = 0.027). Interpretation of the causal associations was very similar to that of the main analysis in both men and women (Supplementary Figures 16-17). The bidirectional DoC model suggested a causal effect of LTPA on BMI in 1981 only in women. In women, adjusting for genetic correlation attenuated the causal association from higher BMI to lower LTPA in 1975 (Models 2-4). The estimation results for the MR-DoC model indicated a positive horizontal pleiotropic effect from PGS BMI to LTPA among women in 1981 and 1990, suggesting that a higher PGS BMI was associated with a higher LTPA independent of BMI in those years (Model 4, see also Supplementary Figures 18-19).

## 4 Discussion

We triangulated evidence for the bidirectional causal relationships between LTPA and BMI using longitudinal, MR and twin models. Across all three approaches, we consistently observed a causal effect of higher BMI on lower LTPA, particularly in the later stages of the follow-up. This finding may highlight the role of high BMI in limiting LTPA in older age, but it may also reflect period effects related to environmental changes over decades. The results regarding the effect of LTPA on BMI were less conclusive, which could likely be explained, at least in part, by the long lags between the measurement points and the weakness of the genetic instruments for LTPA.

### 4.1 Within-person cross-lagged effects between BMI and LTPA (RI-CLPM)

Our RI-CLPM utilising longitudinal individual-level data indicated that a greater deviation in the person’s own BMI level in 1981 and 1990 prospectively predicted a decrease in that person’s own LTPA level 10 and 20 years later. We are not aware of any previous studies that have used any form of cross-lagged panel model to investigate bidirectional associations between PA or LTPA and BMI in adulthood. Some studies that used regression models to investigate the prospective associations in both directions concluded that higher BMI seems to lead to lower PA, but not vice versa.^8,19,66,67^ The lack of evidence for a prospective effect of PA on BMI may be due to the excessively long time lags between measurements and a dilution of the effect.^8^ While PA may support short-term weight management, sustaining an active lifestyle over the long term can be challenging,^68^ as evidenced by the low correlations between LTPA over time in our analysis. In our RI-CLPMs, the observation of time-specific negative residual correlations indicated that, within the given measurement point, a higher-than-usual LTPA and a lower-than-usual BMI tended to co-occur. This pattern may reflect the parallel changes in PA and BMI reported in previous studies,^18,69^ but it may also indicate time-varying confounding due to third variables, such as diet or smoking, which were not controlled for in our analysis.^70^

### 4.2 Evidence for causal relationships between LTPA and BMI based on MR

Our MR findings indicated that a higher BMI causally associates with a lower LTPA, especially at the later stages of follow-up. In adults, the existing published evidence is mainly limited to two-sample MR studies.^33–37^ Studies using summary statistics derived from GWAS of device-based PA data in the UK biobank^26,35^ have suggested a bidirectional negative causal relationship between PA at various intensities (overall, moderate and vigorous) and BMI.^33,35^ However, horizontal pleiotropy has been reported a confounder, especially of the causal effect of BMI on PA, in these studies. A recent study found no evidence for a causal effect of higher PA (derived from GWAS of self-reported MVPA, vigorous PA or device-based overall PA in the UK biobank^26^) on lower obesity risk or BMI.^34,36^ A study based on meta-analysis of GWASs of dichotomised self-reported PA, using the largest sample size to date, also suggested a bidirectional causal association,^28,37^ but the causal effect of higher BMI on lower PA was explained by education. The mixed results in these previous two-sample MR studies arose due to the various GWASs used to identify genetic instruments for PA and the various measurement methods, as well as the use of slightly different analysis methods to conduct MR. Moreover, the age-specific or time-specific nature of the causal effects suggested by our findings was not captured by two-sample MR studies.

In our study, the MR results obtained using the PGSs as instruments differed considerably from those based on data summarized from individual SNPs. Using the PGS as an instrument yielded less consistent evidence of causal effects with wide confidence intervals. This is likely because BMI does not provide information on body composition, so the same high BMI can reflect different phenotypes, ranging from high adiposity to high muscularity. Using summarised data from individual SNPs enables consideration of heterogeneity in the effects of BMI-related SNPs on LTPA, which is not possible when a PGS is used as an instrument. The inconsistent evidence for the causal effects of LTPA on BMI is partially due to the poor instruments. We were unable to identify a PGS that worked properly as an instrument for LTPA. Interestingly, a recent study using non-genetic instruments for PA (shocks to PA, such as accidents, and disabilities affecting physical movement) suggested a causal association between higher PA and lower BMI.^71^

### 4.3 Traditional DoC twin models and MR-DoC twin models

We observed a negative genetic correlation between BMI and LTPA, indicating that some genetic factors that predispose individuals to a higher BMI also predispose the to lower LTPA. In the later phases of follow-up, a modest unique environmental correlation was observed, which is a necessary condition for a relationship to be causal.^72^ The bidirectional DoC models suggested causal effects of higher BMI on lower LTPA, but the unidirectional DoC and MR-DoC models that accounted for shared genetic factors suggested relatively consistent causal effects in both directions in the later phases of follow-up.

In women, the estimation results obtained from the MR-DoC models indicated horizontal pleiotropy, as a direct association was evident from higher PGS BMI to higher LTPA, independent of the observed BMI. In this case, a positive horizontal pleiotropic effect, if not taken into account, may lead to an underestimation of the causal effect from higher BMI to lower LTPA. This effect may reflect that women with a higher genetic predisposition to gain weight face greater social pressure to control it, thereby leading them to increase their exercise behaviour independent of their actual BMI. When examining the effects in the opposite direction, we observed that including a weak instrument for LTPA in the MR-DoC models did not affect the causal estimates compared to the traditional unidirectional DoC model.

### 4.4 Strengths and limitations

The main strength of this study was its use of unique longitudinal twin data with repeated measurements from 1975 to 2011, combined with a large sample size, high response rates over time, and genotype information available in a subsample. This enabled us to apply three approaches to examine the plausibility of causal effects from multiple points of view. We explored several PGSs for PA based on recently published GWASs, and LTPA was measured using validated questionnaires.^6^ Our one-sample MR approach enabled an investigation of time- and sex-specific causal relationships between LTPA and BMI, which have rarely been examined in previous MR studies. However, several limitations exist. One is that BMI does not provide information on body composition. Another is that self-reported LTPA is susceptible to recall and social-desirability biases. Moreover, the LTPA questionnaire was slightly different in 1990, and a considerable proportion of the participants did not have measured LTPA at all four measurement points. Lastly, we were unable to identify a PGS that could serve as a valid genetic instrument for LTPA, so reliable MR-based causal inference was limited.

### 4.5 Conclusion

Different analytical methods may produce conflicting results regarding causal effects, which may also vary by age or time. Our findings suggest a causal association between higher BMI and lower LTPA, particularly at the later stages of follow-up. This highlights the role of high BMI in limiting LTPA in older age, while also emphasising the importance of weight management in maintaining a physically active lifestyle later in life. We found no conclusive evidence for a causal effect of higher LTPA on lower BMI, which may challenge the effectiveness of LTPA in weight management.

## Data Availability

Data are available to authorized researchers with institutional review board/ethics approval and an institutionally approved study plan. Overall cohort data are available through the Institute for Molecular Medicine Finland (FIMM) Data Access Committee (DAC). For more details, please contact the FIMM DAC. Data of participants with measured genetic information are available through THL Biobank (see the THL Biobank website https://thl.fi/en/research-and-development/thl-biobank/for-researchers).

## 5 Declarations

## Acknowledgments

We thank all the FTC study participants, the FTC research team members, and the persons involved in the data collection. We thank Monica Madore from Scribendi (www.scribendi.com) for editing a draft of this manuscript.

## Author contributions

AK conceptualised the research question and study design, performed the statistical analyses, and drafted the first version of the manuscript. LJ conceptualised the research question and study design, participated in drafting the first version of the manuscript, and calculated the genome-wide PRS. UE contributed to the the research question and to critical interpretation of the findings. AP, KW, MO and SA contributed to the critical interpretation of the findings. TP contributed to the FTC data management and counselling on the genetic data. JK contributed to the data collection of the FTC and significantly to the study design and writing. ES supervised the study, conceptualised the research question and study design, contributed significantly to the writing and acquired funding for the study. All authors contributed to revising and editing the manuscript. All authors have read and approved the final version of the manuscript and agree with the order of presentation of the authors.

## Informed consent

All participants provided informed consent, when replying to questionnaires. Blood samples for DNA analyses were collected during in-person clinical studies after written informed consent was signed.

