## Supplementary material for "The causal relationships between leisure-time physical activity and body mass index in adulthood: A triangulation study"

Kankaanpää et al.

Figures:

Tables:

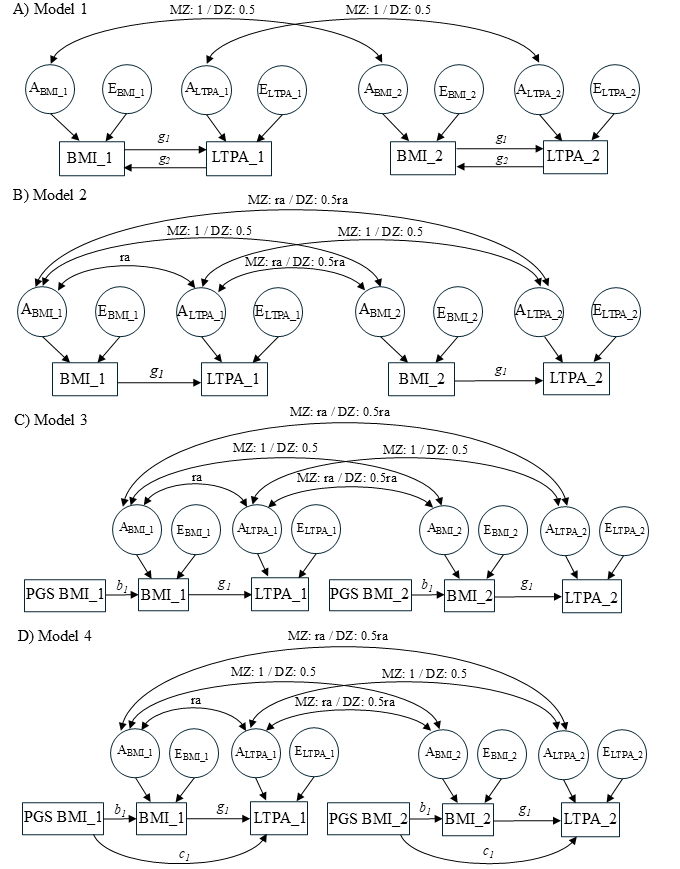

**Supplementary Figure 1.** Path diagrams of the direction of causation (DoC) and Mendelian Randomisation (MR)-DoC twin models: A) bidirectional DoC model, B) unidirectional DoC model adjusted for genetic correlation, C) MR-DoC model, pleiotropic path constrained to zero, and D) MR-DoC model, pleiotropic path c estimated.

Note. Double-headed arrows represent genetic correlations. ra, cross-trait additive genetic correlation. Models B‒D were also applied in the reverse direction to estimate the causal effect of LTPA on BMI.

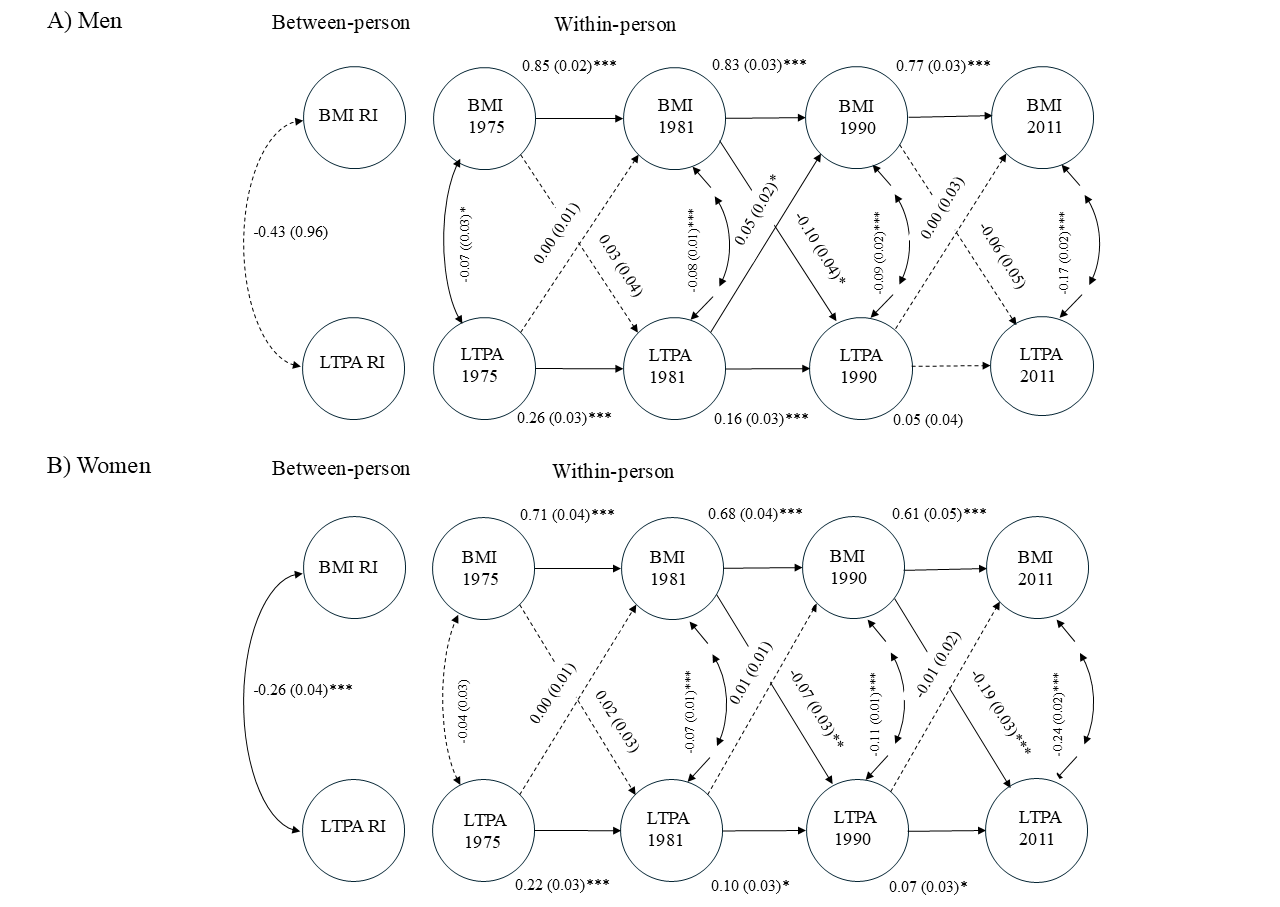

**Supplementary Figure 2.** Estimation results of the random-intercept cross-lagged panel models (RI-CLPMs) in A) men (n = 11,282) and B) women (n = 11,414).

The circles denote latent variables. Standardised estimates (standard errors) are presented. ****p* < 0.001, ***p* < 0.01, * *p* < 0.05*.*

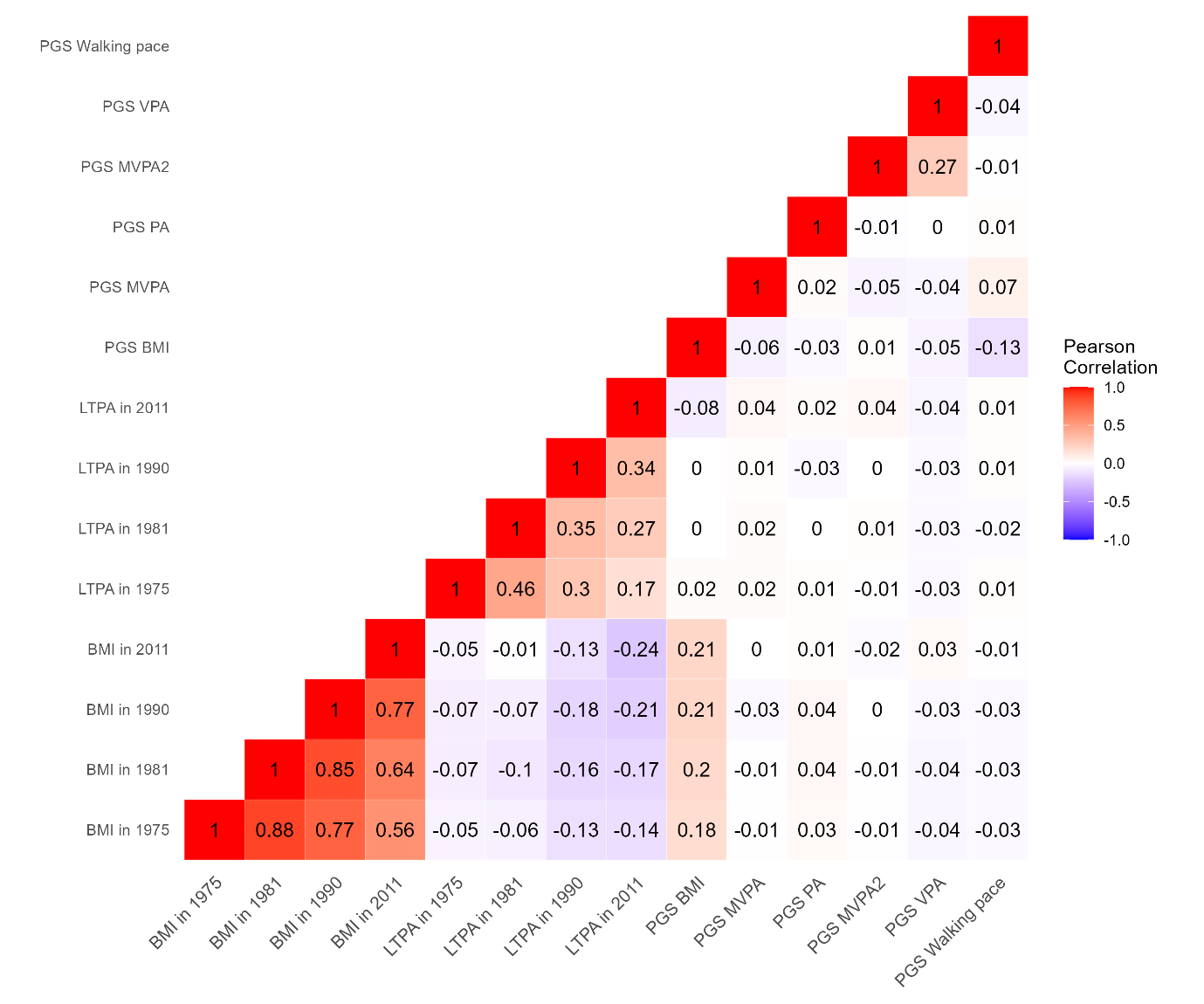

**Supplementary Figure 3.** The Pearson’s correlation coefficients among the study variables in subsample having measured genetic data (n = 8,527). Pairwise complete observations were used.

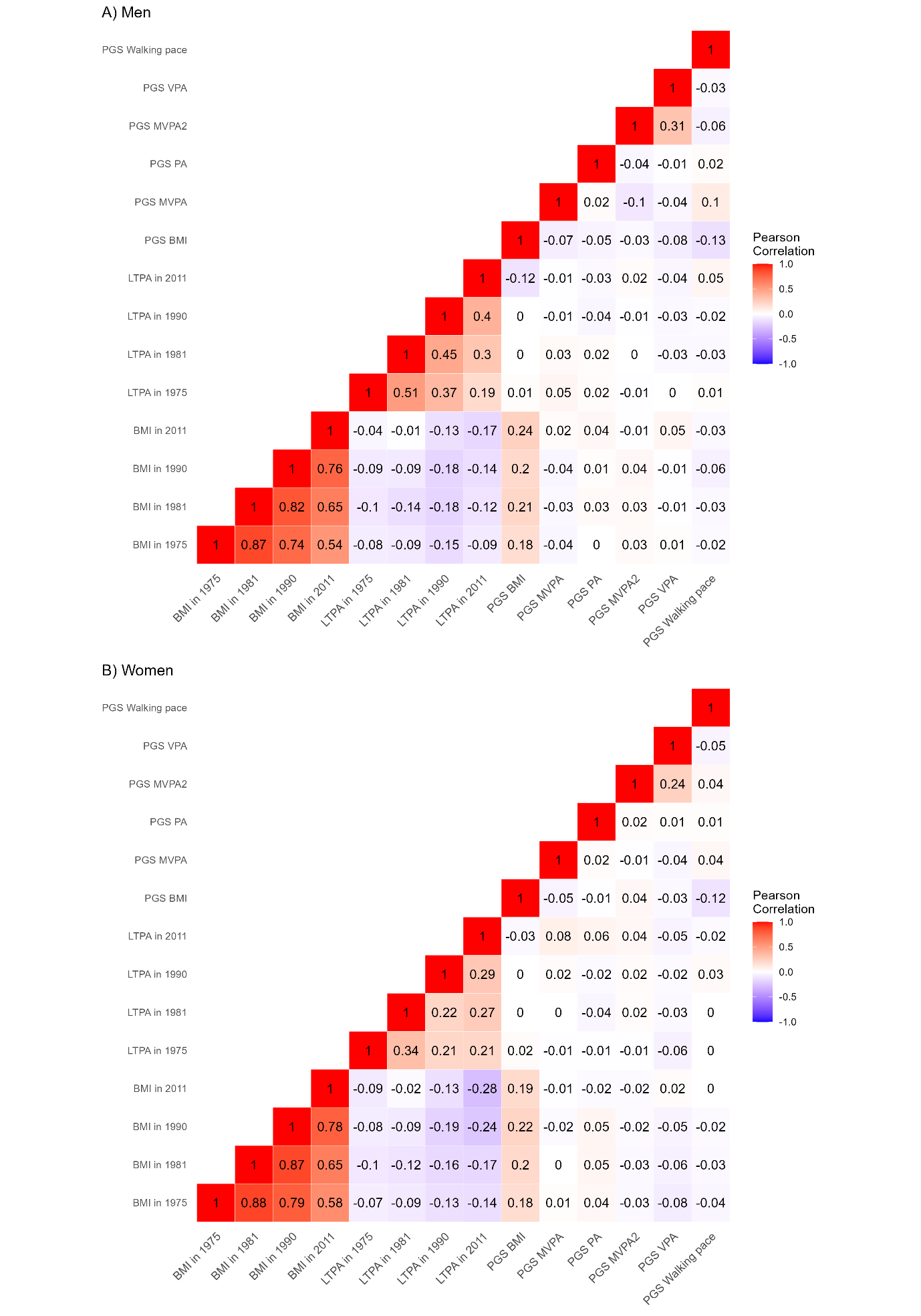

**Supplementary Figure 4.** The Pearson’s correlations among the study variables in A) men (n = 4,062) and B) women (n = 4,465).

Pairwise complete observations were used.

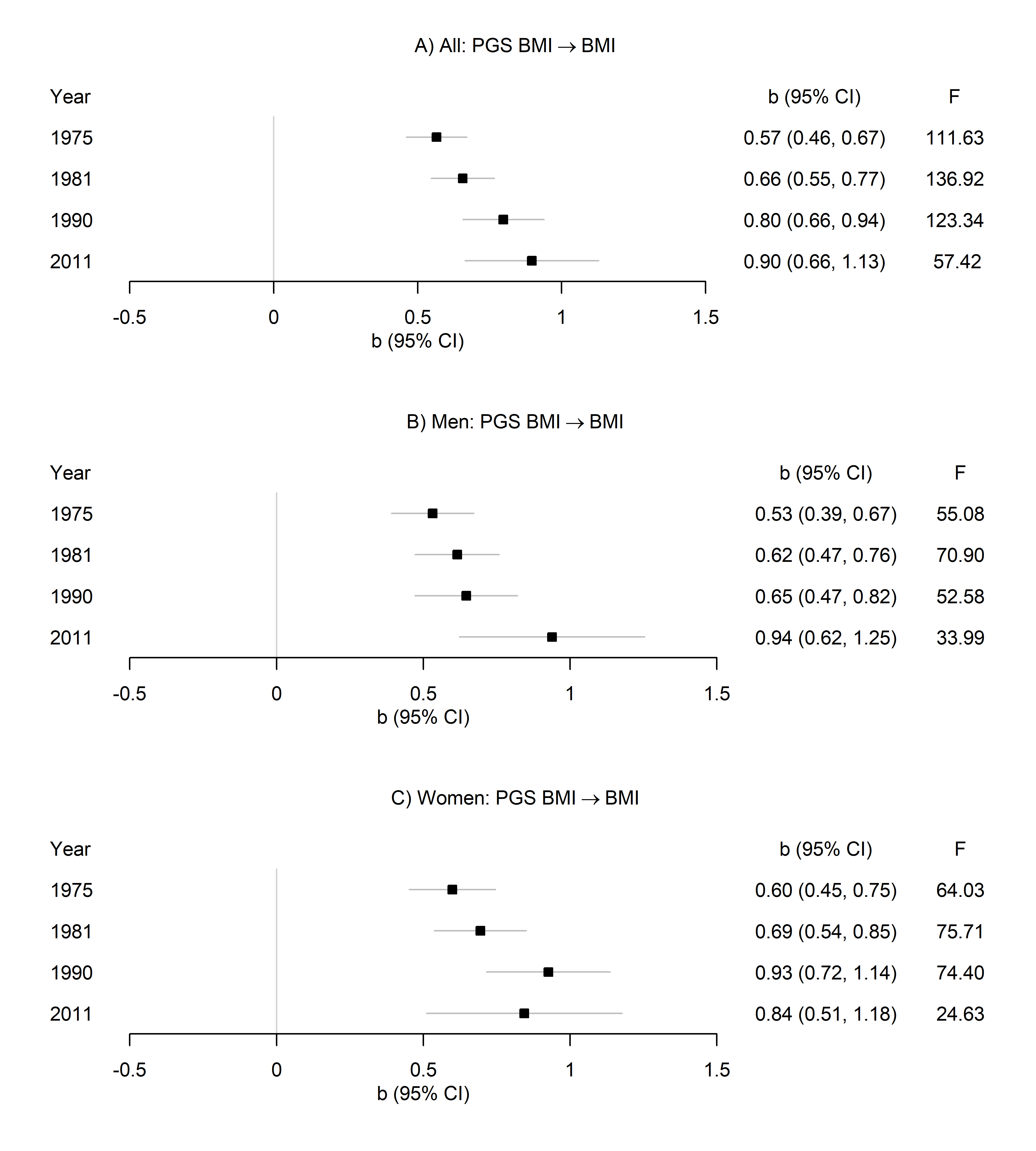

**Supplementary Figure 5.** The associations between the polygenic score for body mass index (PGS BMI) with BMI in A) all, B) men and C) women.

Unstandardised regression coefficients (95% Confidence Intervals) and F statistics are presented. Sample sizes at the four time points were A) all participants (n = 3,513; 3,452; 2,744 and 1,247), B) men (n = 1,609; 1,573; 1,224 and 552) and C) women (n = 1,904; 1,879; 1,520 and 695).

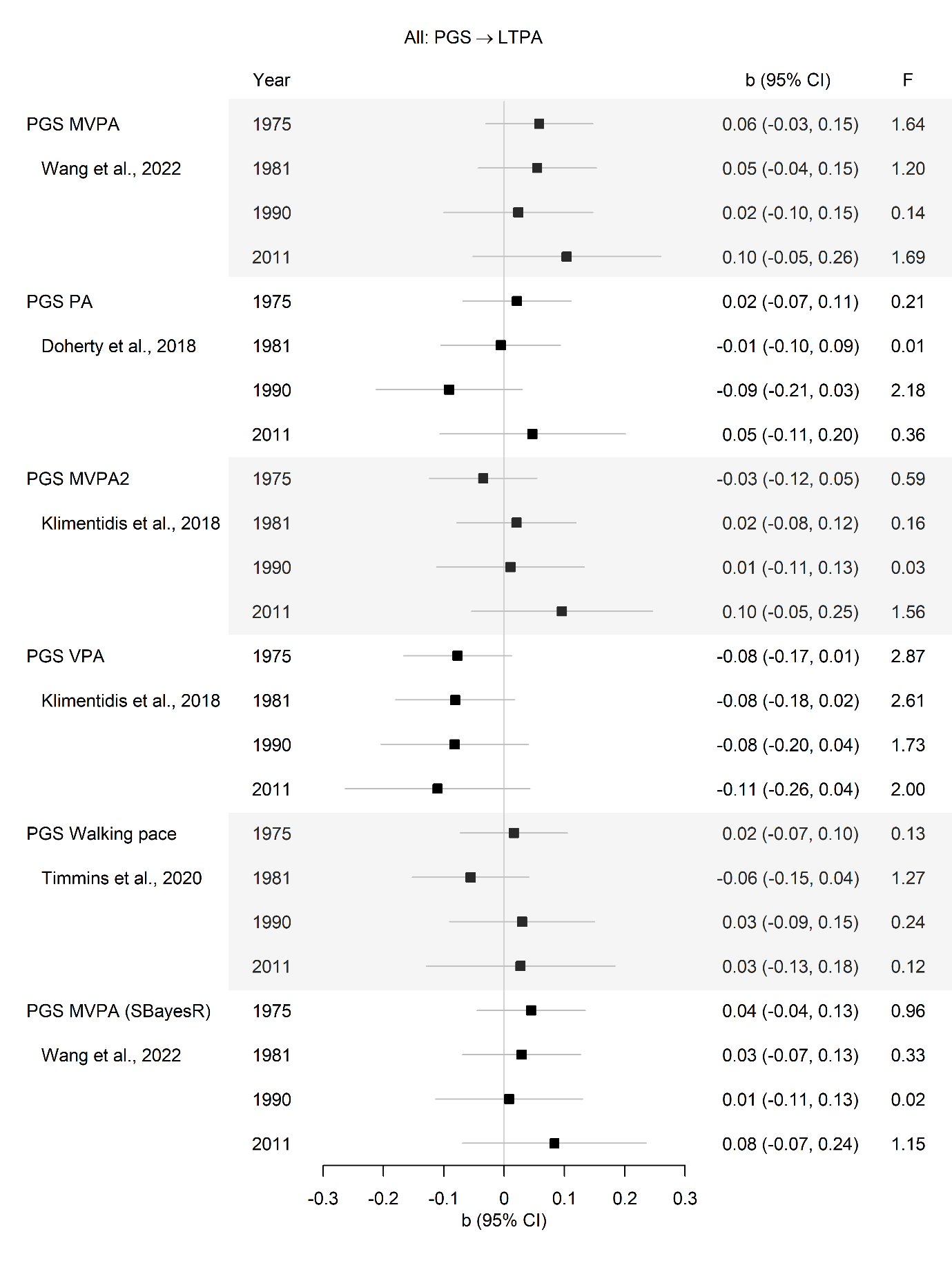

**Supplementary Figure 6.** The associations between the polygenic scores (PGSs) with leisure-time physical activity (LTPA) in all participants.

Sample sizes at the four time points were n = 3,525; 3,472; 2,730 and 1,160. Unstandardised regression coefficients (95% Confidence Intervals) and F statistics are presented. SBaysR, genome-wide PGS.

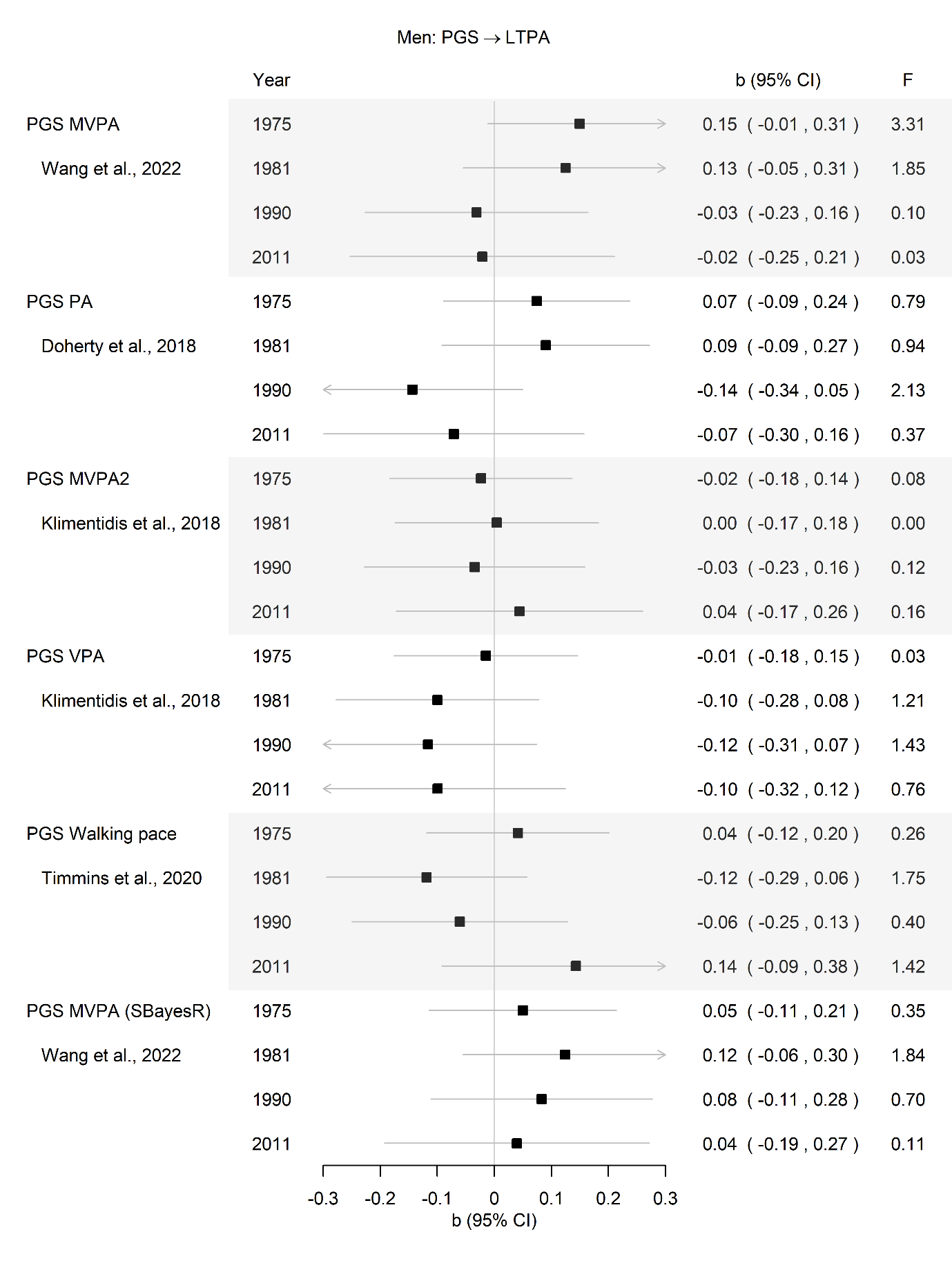

**Supplementary Figure 7.** The associations between the polygenic scores (PGSs) with leisure-time physical activity (LTPA) in men.

Sample sizes at the four time points were n = 1,607; 1,578; 1,223 and 522. Unstandardised regression coefficients (95% Confidence Intervals) and F statistics are presented.

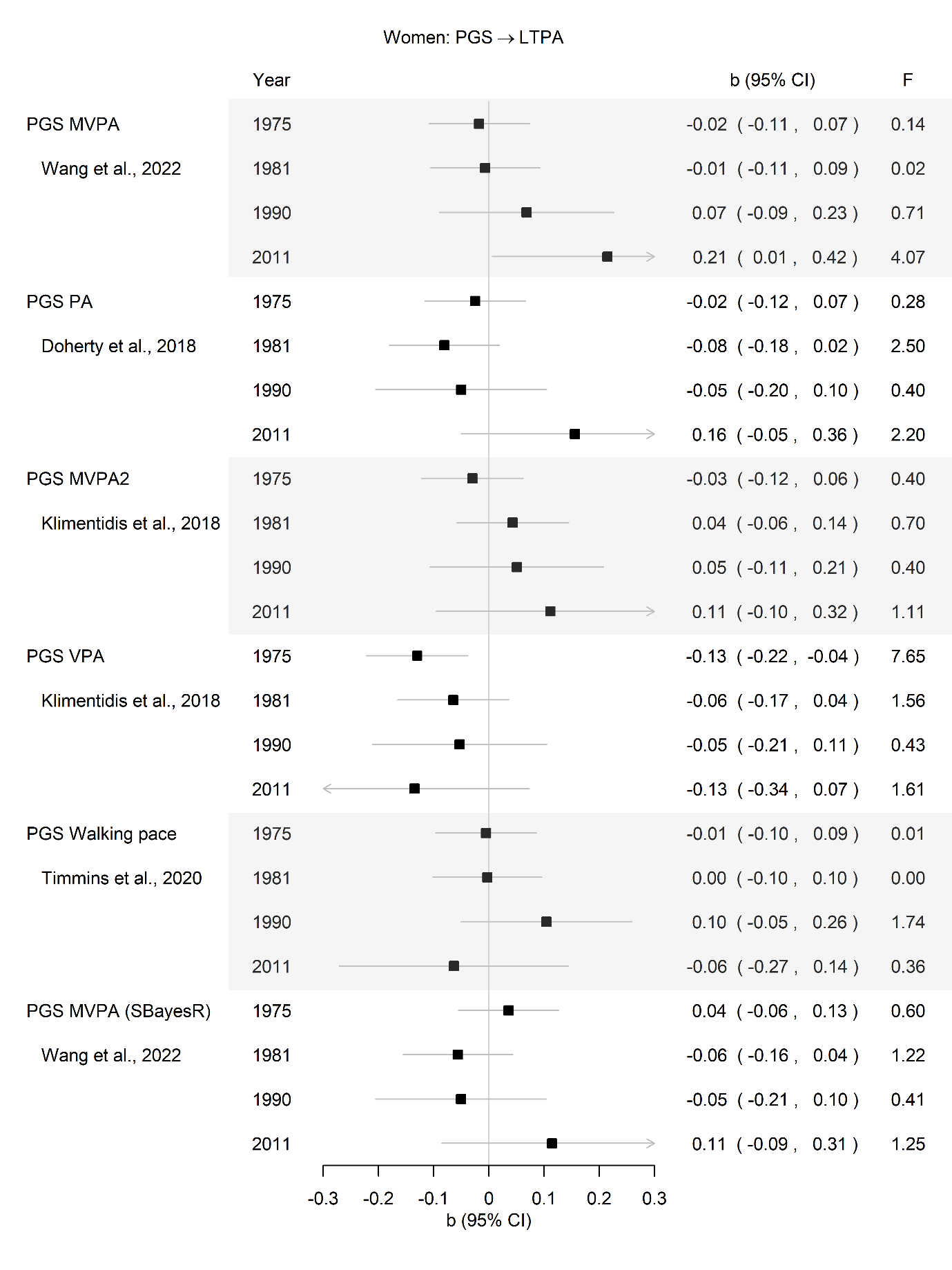

**Supplementary Figure 8.** The associations between the polygenic scores (PGSs) with leisure-time physical activity (LTPA) in women.

Sample sizes at the four time points were n = 1,918; 1,894; 1,507 and 638. Unstandardised regression coefficients (95% Confidence Intervals) and F statistics are presented.

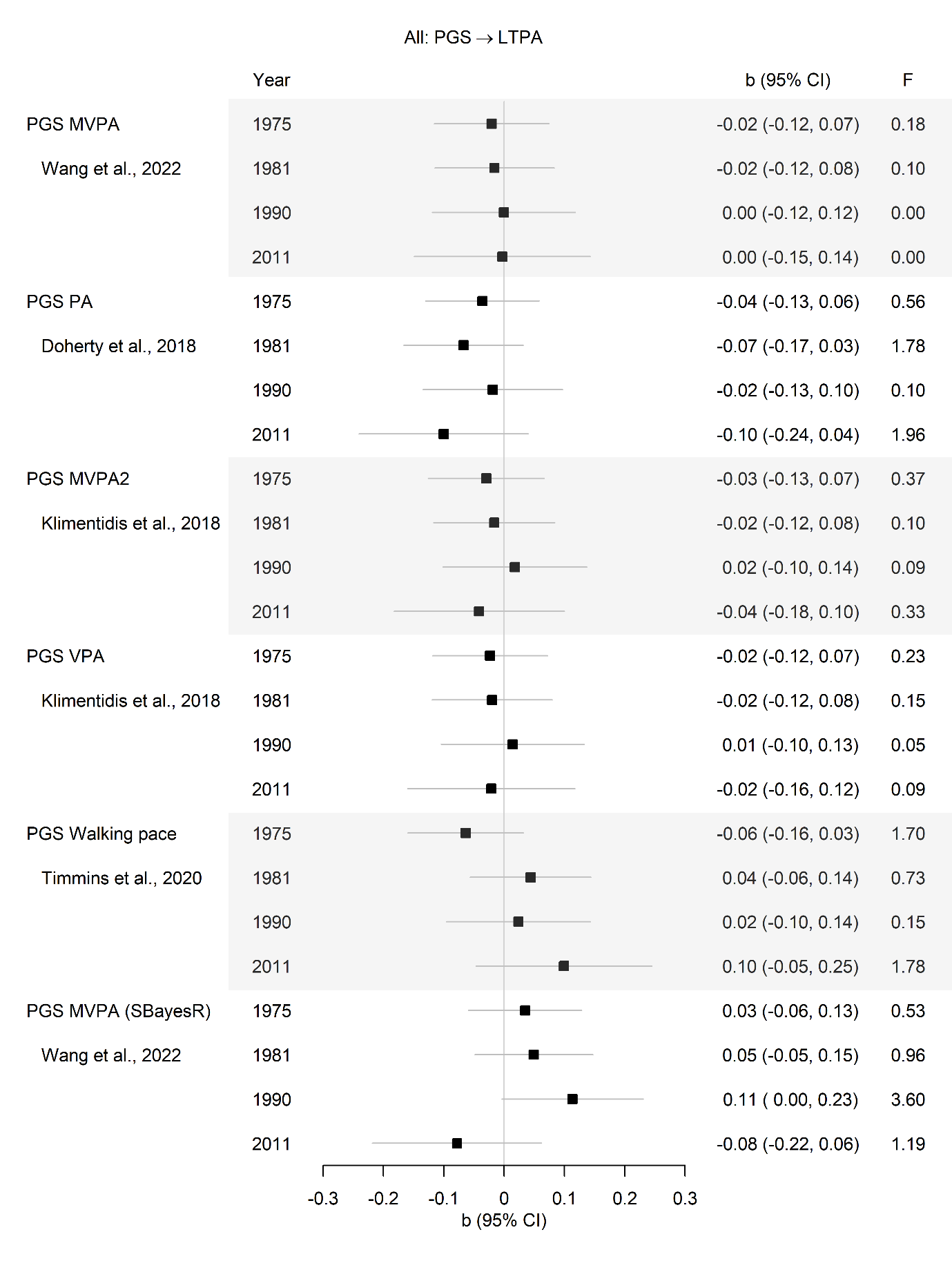

**Supplementary Figure 9.** Replication using data from twin 2: The associations between the polygenic scores (PGSs) with leisure-time physical activity (LTPA) in all participants.

Sample sizes at the four time points were n = 3,546; 3,435; 2,736 and 1,186. Unstandardised regression coefficients (95% Confidence Intervals) and F statistics are presented. SBaysR, genome-wide PGS.

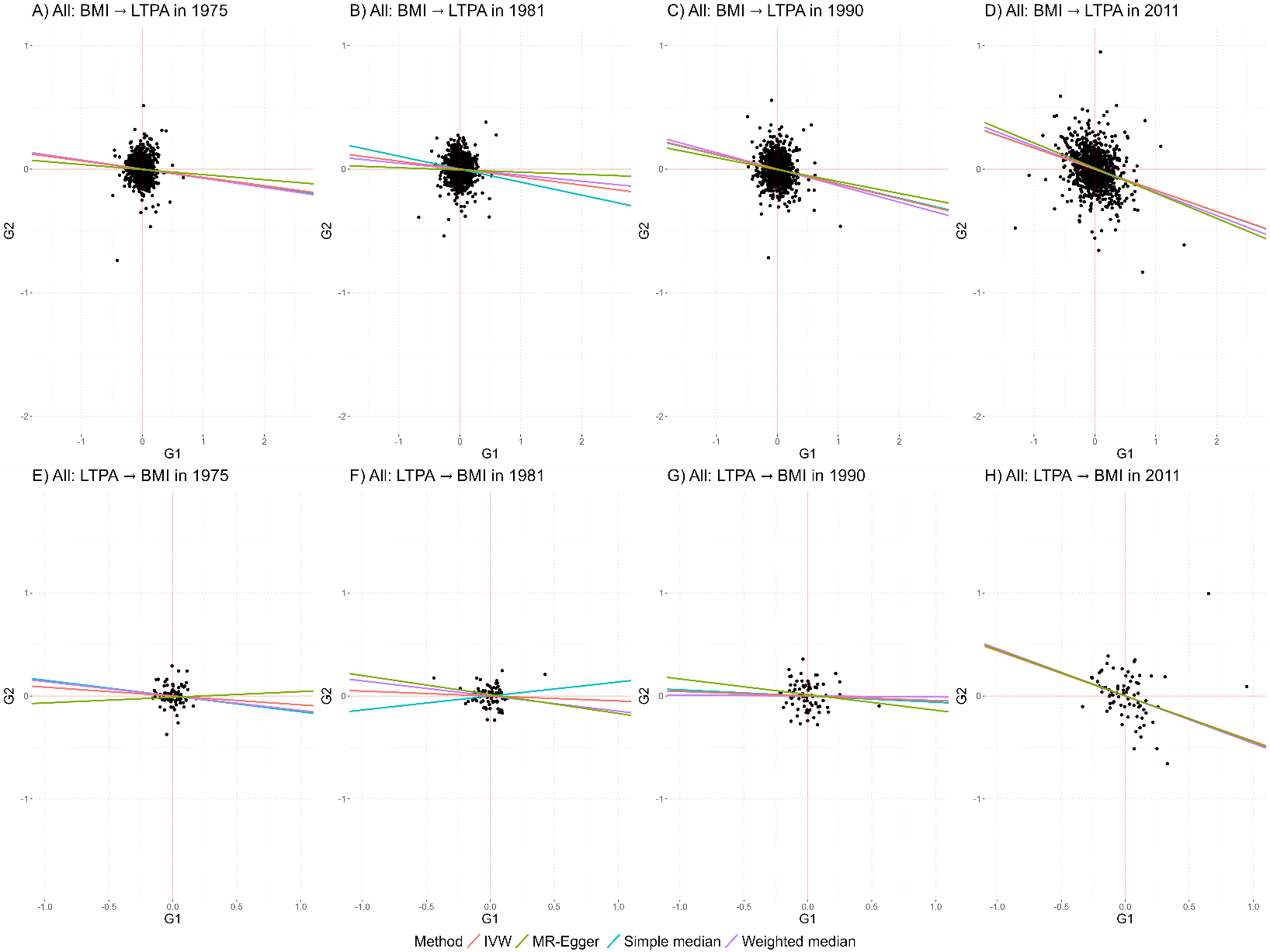

**Supplementary Figure 10.** Scatter plot illustrating the associations of SNPs included in the polygenic score for exposure with measured exposure (G1, x axis) and measured outcome (G2, y axis).

The slopes of the lines correspond to the causal estimates of BMI on LTPA (A‒D) and LTPA on BMI (E‒H) assessed using different Mendelian Randomisation (MR) methods. IVW, inverse variance weighted.

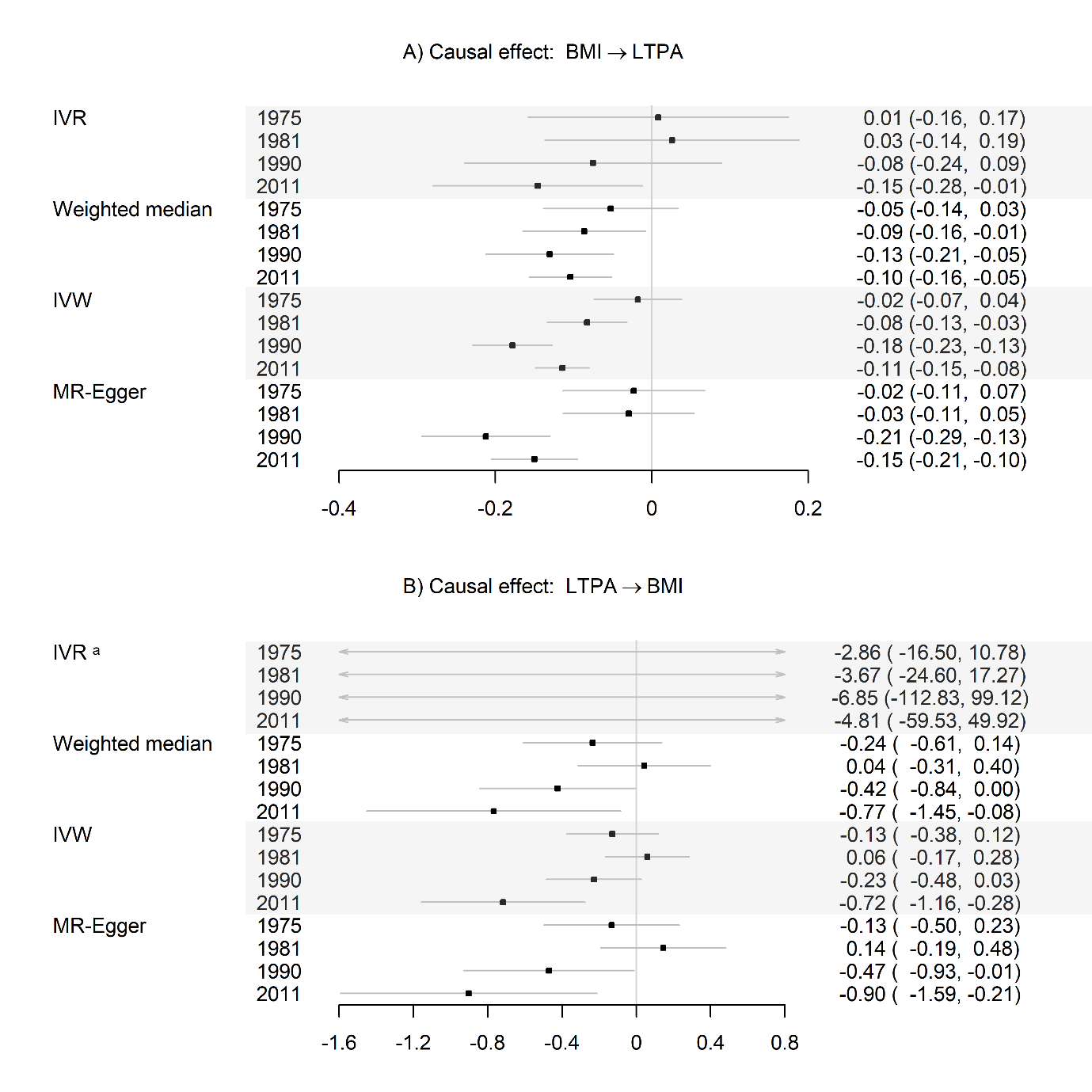

**Supplementary Figure 11.** Replication using data from twin 2: Causal effects (95% confidence intervals) of A) body mass index (BMI) on leisure-time physical activity (LTPA) and B) LTPA on BMI, estimated using several Mendelian randomisation (MR) methods.

IVR, Instrumental Variable Regression; IVW, Inverse Variance Weighted. Unstandardised coefficients are presented. Sample sizes at the four time points were: 3,517; 3,416; 2,715 and 1,164, respectively. Note. a PGS MVPA was a poor instrument for LTPA.

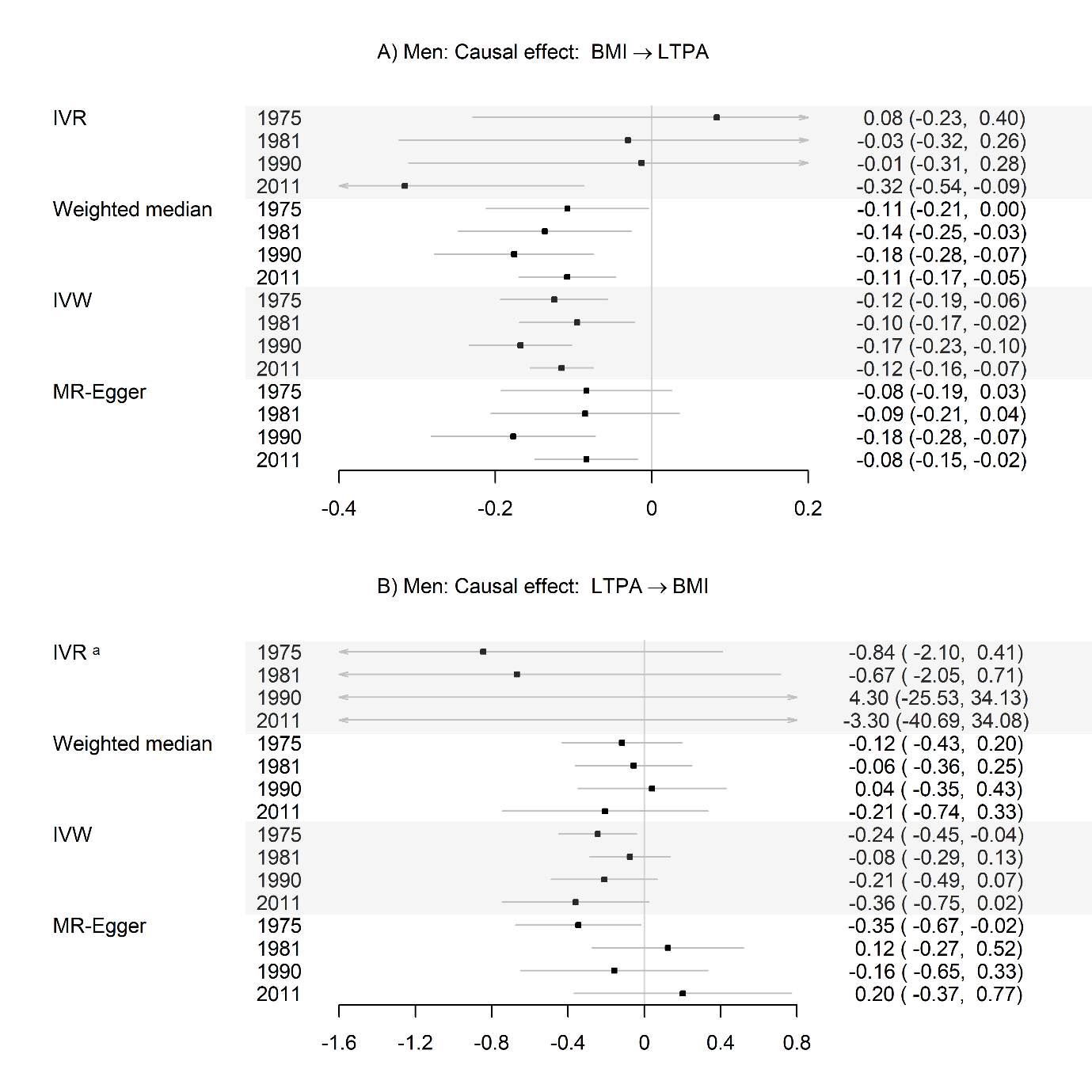

**Supplementary Figure 12.** Sex-stratified analysis: Causal effects (95% confidence intervals) of A) body mass index (BMI) on leisure-time physical activity (LTPA) and B) LTPA on BMI in men, estimated using several Mendelian randomisation (MR) methods.

IVR, Instrumental Variable Regression; IVW, Inverse Variance Weighted. Unstandardised coefficients are presented. Sample sizes at the four time points were: 1,603; 1,569; 1,217 and 506, respectively. Note. a PGS MVPA was a poor instrument for LTPA.

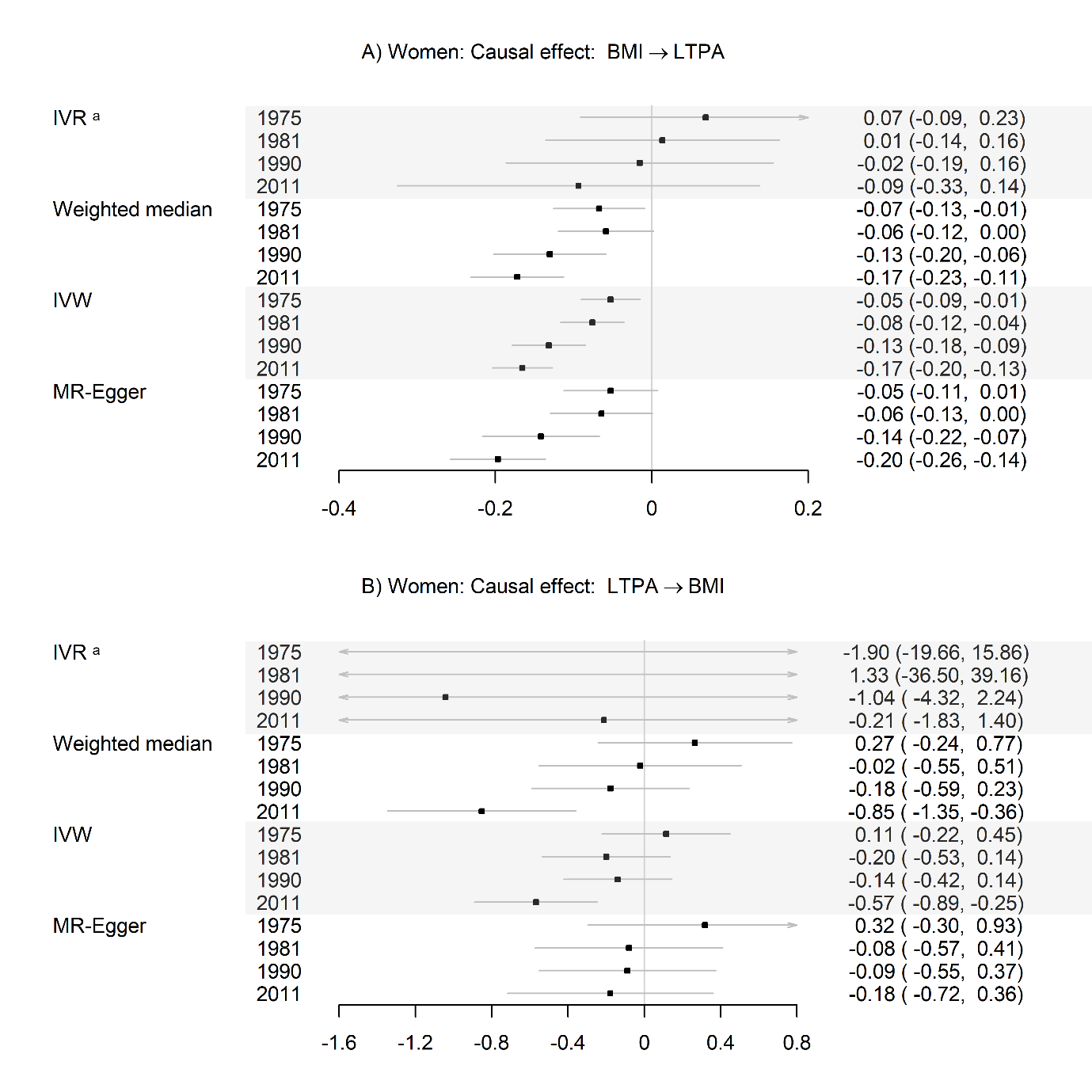

**Supplementary Figure 13.** Sex-stratified analysis: Causal effects (95% confidence intervals) of A) body mass index (BMI) on leisure-time physical activity (LTPA) and B) LTPA on BMI in women, estimated using several Mendelian randomisation (MR) methods.

IVR, Instrumental Variable Regression; IVW, Inverse Variance Weighted. Unstandardised coefficients are presented. Sample sizes at the four time points were: 1,767; 1,742; 1,404 and 591, respectively. Note. ^a^ PGS MVPA was a poor instrument for LTPA.

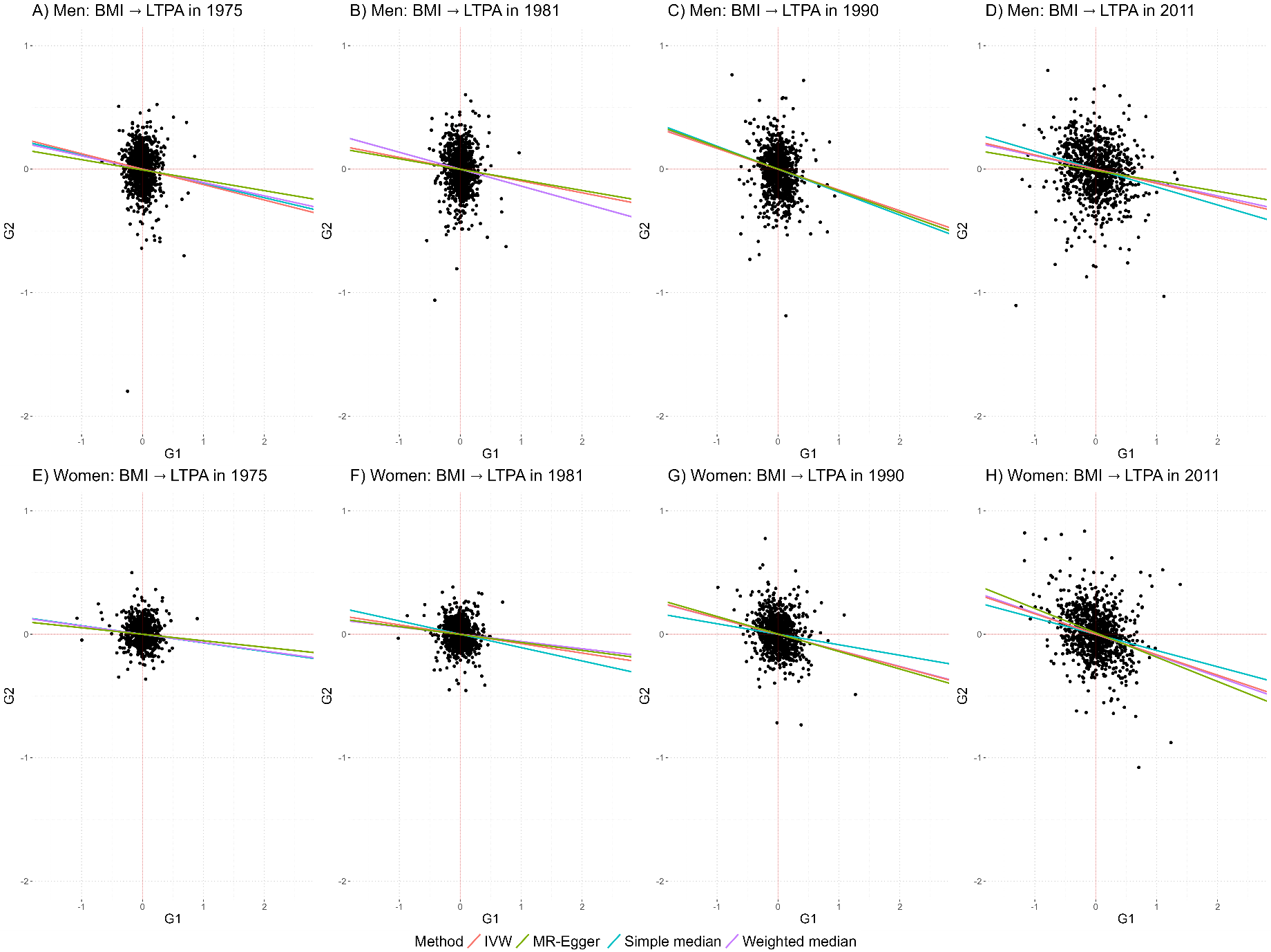

**Supplementary Figure 14.** Sex-stratified analysis: Scatter plot illustrating the associations of SNPs included in the polygenic score for body mass index (PGS BMI) with BMI (G1, x axis) and leisure-time physical activity (LTPA) (G2, y axis).

The slopes of the lines correspond to the causal estimates of BMI on LTPA assessed using different Mendelian Randomisation (MR) methods in men (A‒D), and women (E‒H). IVW, inverse variance weighted.

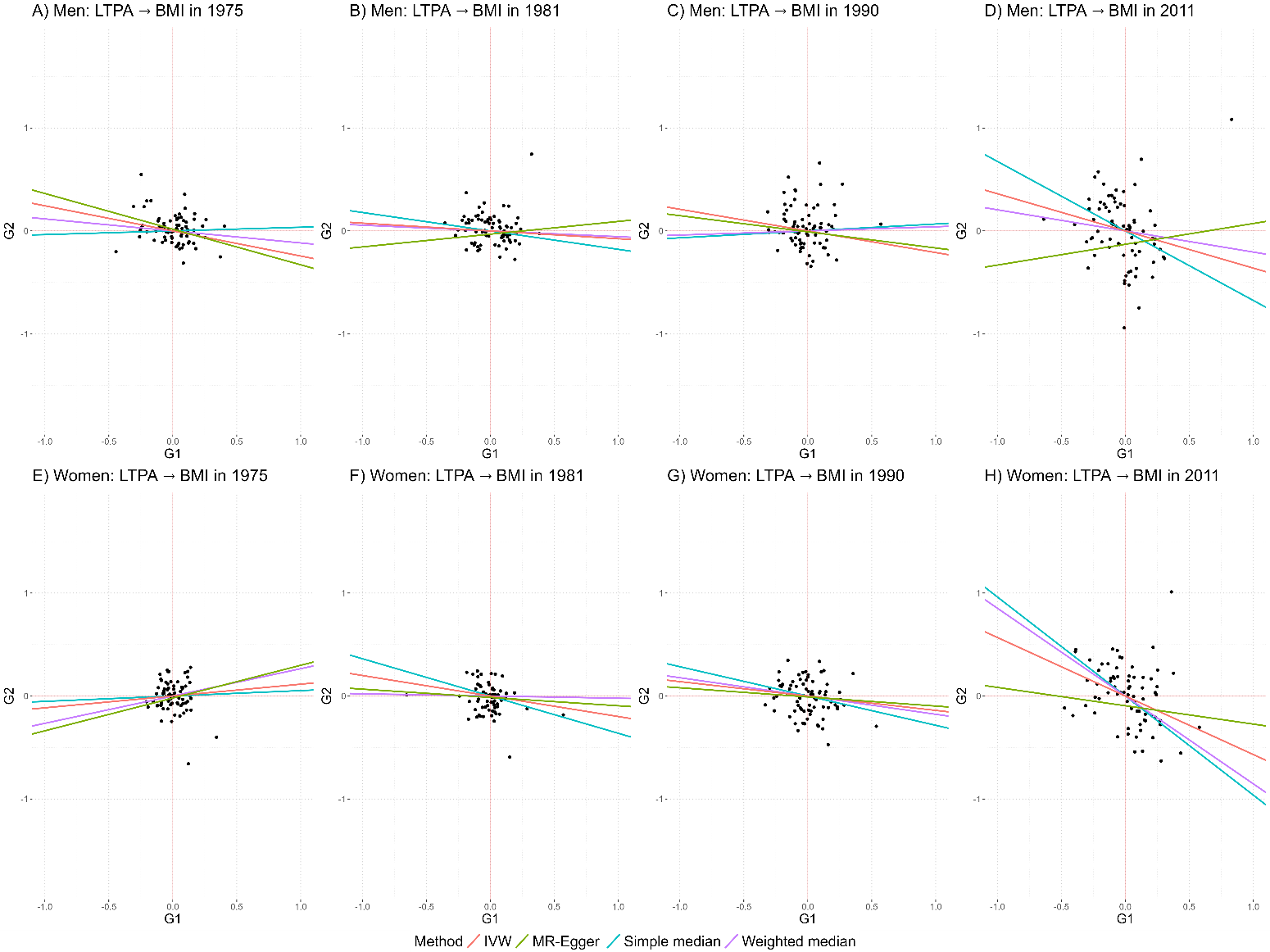

**Supplementary Figure 15.** Sex-stratified analysis: Scatter plot illustrating the associations of SNPs included in the polygenic score for moderate-to-vigorous physical activity MVPA (PGS MVPA) with leisure-time physical activity (LTPA) (G1, x axis) and body mass index (BMI) (G2, y axis).

The slopes of the lines correspond to the causal estimates of LTPA on BMI assessed using different Mendelian Randomisation (MR) methods in men (A‒D), and women (E‒H). IVW, inverse variance weighted.

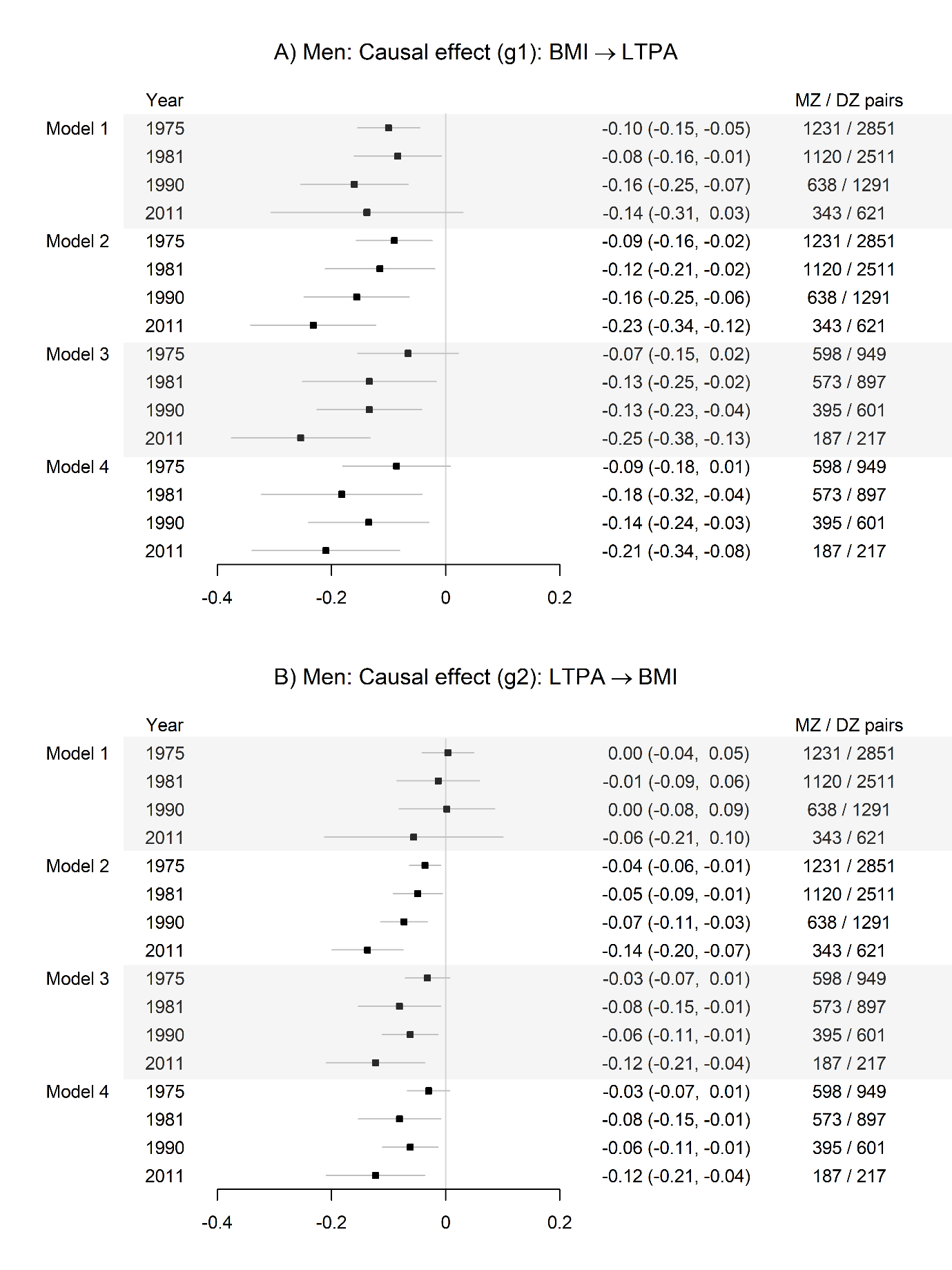

**Supplementary Figure 16.** Sex-stratified analysis: Causal effects of A) body mass index (BMI) on leisure-time physical activity (LTPA) and B) LTPA on BMI in years 1975, 1981, 1990 and 2011 assessed using Direction of Causation (DoC) and Mendelian Randomisation (MR)-DoC twin models in men.

Standardised coefficients (95% confidence intervals) are presented. MZ, monozygotic; DZ, dizygotic.

Model 1, bidirectional DoC twin model.

Model 2, unidirectional DoC model adjusted for genetic correlation.

Model 3, MR-DoC model, no horizontal pleiotropy assumed.

Model 4, MR-DoC model, horizontal pleiotropy estimated.

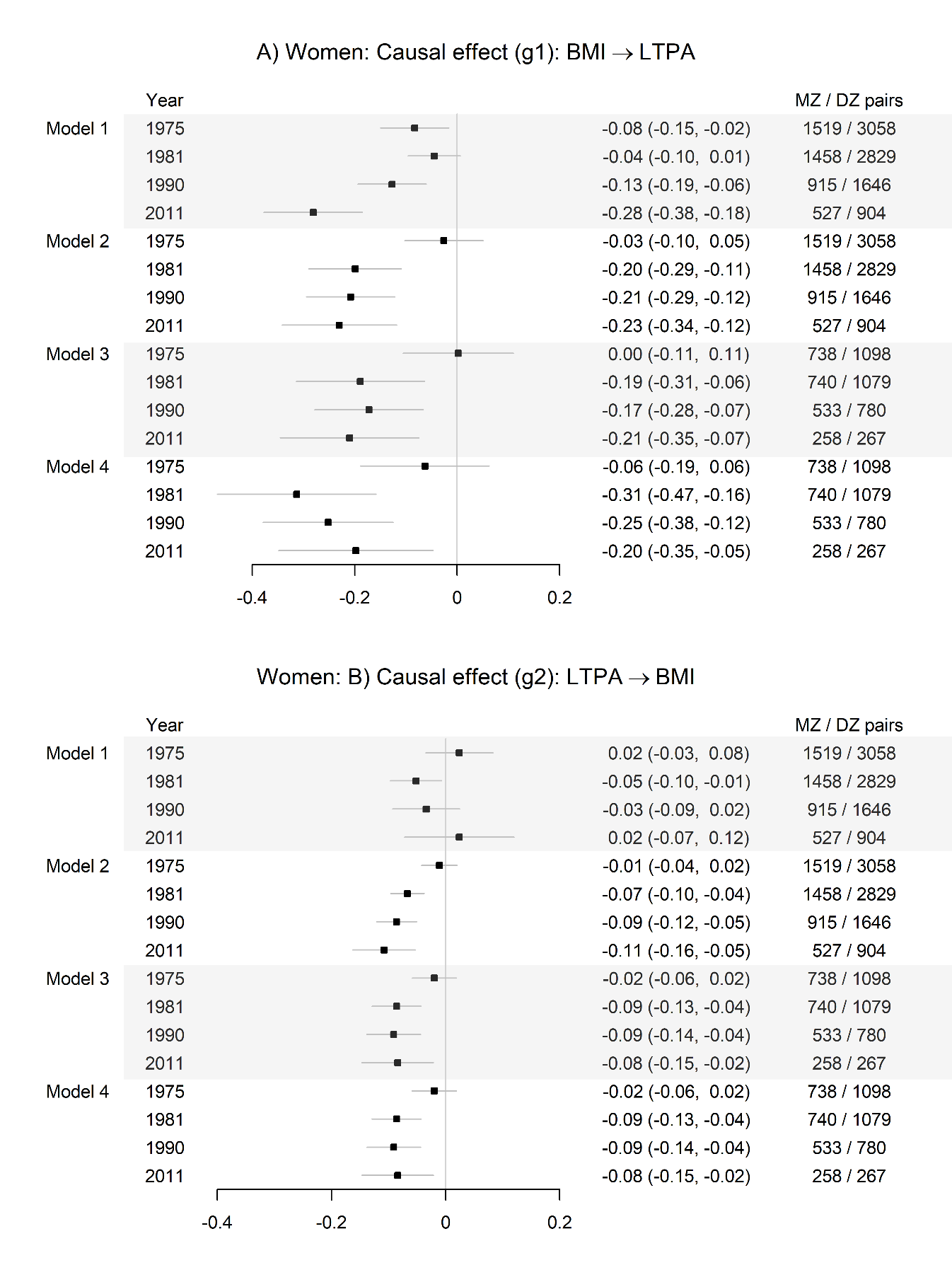

**Supplementary Figure 17.** Sex-stratified analysis: Causal effects of A) body mass index (BMI) on leisure-time physical activity (LTPA) and B) LTPA on BMI in years 1975, 1981, 1990 and 2011 assessed using Direction of Causation (DoC) and Mendelian Randomisation (MR)-DoC twin models in women.

Standardised coefficients (95% confidence intervals) are presented. MZ, monozygotic; DZ, dizygotic.

Model 1, bidirectional DoC twin model.

Model 2, unidirectional DoC model adjusted for genetic correlation.

Model 3, MR-DoC model, no horizontal pleiotropy assumed.

Model 4, MR-DoC model, horizontal pleiotropy estimated.

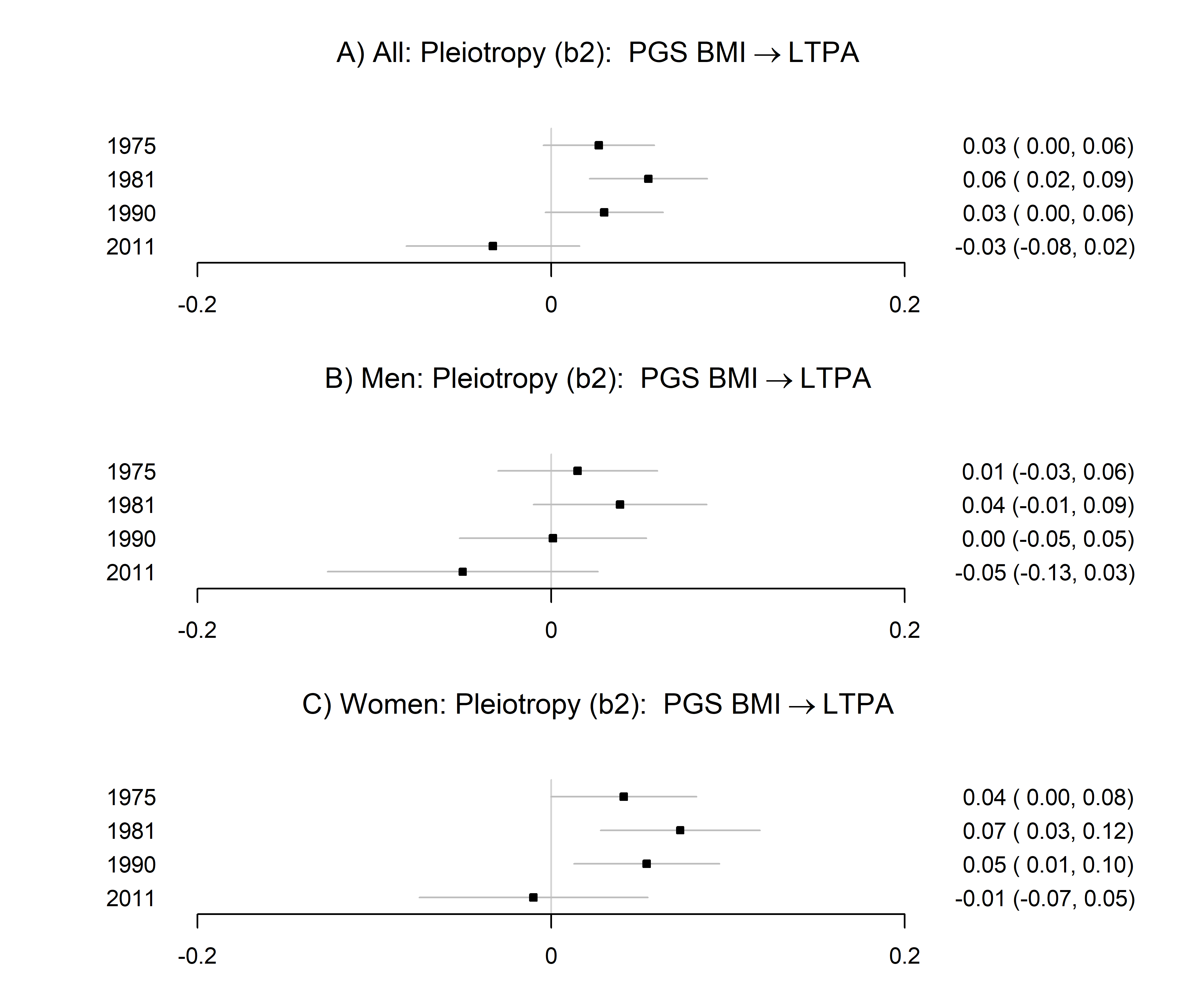

**Supplementary Figure 18.** Estimated horizontal pleiotropy from polygenic score for body mass index (PGS BMI) to leisure-time physical activity (LTPA) estimated using MR-DoC model in A) all, B) men and C) women. Standardised regression coefficients (95% confidence intervals) are presented.

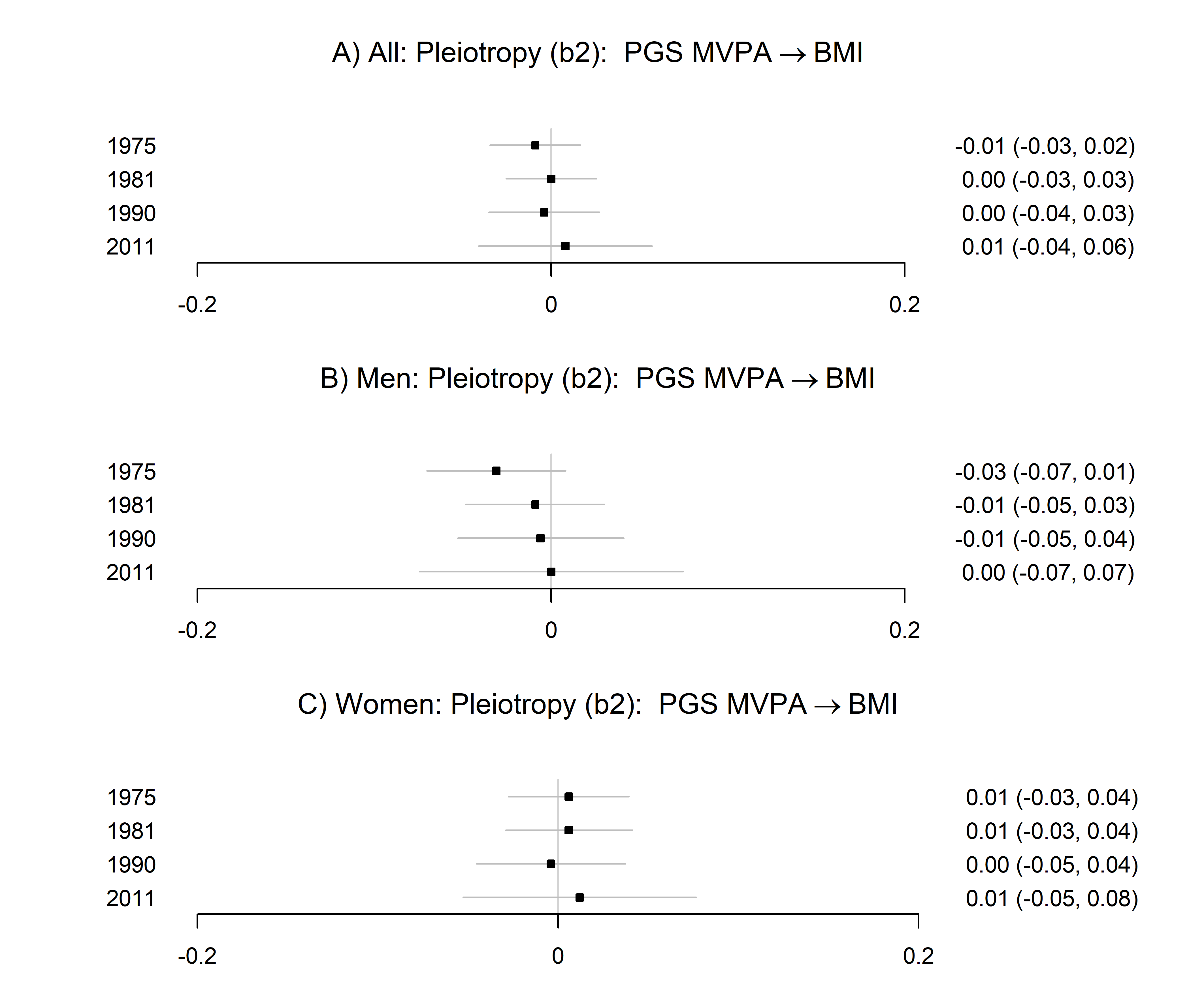

**Supplementary Figure 19.** Estimated horizontal pleiotropy from polygenic score for moderate-to-vigorous physical activity (PGS MVPA) to body mass index (BMI) estimated using MR-DoC model in A) all, B) men and C) women. Standardised regression coefficients (95% confidence intervals) are presented.

**Supplementary Table 1.** The candidate instrumental variables for LTPA.

| Polygenic score | Reference | Outcome | Number of SNPs ^a^ |
| --- | --- | --- | --- |
| PGS MVPA | Wang et al., 2022 | Self-reported MVPA | 76 ^b^ |
| PGS PA | Doherty et al., 2018 | Accelerometer assessed overall PA | 2 |
| PGS MVPA2 | Klimentidis et al., 2018 | Self-reported MVPA | 8 |
| PGS VPA | Klimentidis et al., 2018 | Self-reported VPA | 5 |
| PGS Walking pace | Timmins et al., 2020 | Walking pace | 65 |
| MVPA, moderate-to-vigorous-physical activity; VPA, vigorous physical activity.  ^a^ Number of SNPs included in the PGS. SNPs were also limited to have a minor allele frequency greater than 1% and imputation quality (INFO score) better than 0.8.  ^b^ The included SNPs were those that were significant either for sedentary time or MVPA, or both. | | | |

**Supplementary Table 2.** Sex-stratified analysis: Descriptive statistics of the study variables in full cohort and subsample of twin individuals having measured genetic data available.

|  | Full cohort | | | | Subsample with genetic data | | | |
| --- | --- | --- | --- | --- | --- | --- | --- | --- |
|  | Men (n = 11,282) | | Women (n = 11,414) | | Men (n = 4,062) | | Women (n = 4,465) | |
|  | n | Mean (SD) or % | n | Mean (SD) or % | n | Mean (SD) or % | n | Mean (SD) or % |
| Zygosity | 11,282 |  | 11,414 |  | 4,062 |  | 4,465 |  |
| Unsure | 1,021 | 9.0 | 858 | 7.5 | - |  | - |  |
| Monozygotic | 2,987 | 26.5 | 3,462 | 30.3 | 1,295 | 31.9 | 1,595 | 35.7 |
| Dizygotic | 7,274 | 64.5 | 7,094 | 62.2 | 2,767 | 68.1 | 2,870 | 64.3 |
| Age at baseline |  |  |  |  |  |  |  |  |
| in 1975 | 11,282 | 30.5 (8.9) | 11,414 | 30.1 (8.5) | 4,062 | 32.9 (8.4) | 4,310 | 33.0 (8.7) |
| in 1981 | 9,956 | 36.7 (8.9) | 10,506 | 36.2 (8.6) | 3,806 | 39.0 (8.4) | 4,140 | 39.2 (8.8) |
| in 1990 | 5,692 | 44.8 (7.7) | 6,801 | 44.2 (7.8) | 2,807 | 47.4 (7.7) | 3,284 | 47.1 (7.8) |
| in 2011 | 3,699 | 60.4 (3.7) | 4,633 | 60.1 (3.7) | 1,277 | 60.6 (3.7) | 1,478 | 60.4 (3.7) |
| Body mass index (kg/m^2^) | |  |  |  |  |  |  |  |
| in 1975 | 10,287 | 23.6 (3.0) | 10,556 | 22.0 (3.6) | 3,898 | 24.0 (2.9) | 4,151 | 22.5 (3.2) |
| in 1981 | 9,768 | 24.4 (3.0) | 10,306 | 22.7 (3.7) | 3,762 | 24.7 (3.0) | 4,087 | 23.3 (3.5) |
| in 1990 | 5,660 | 25.4 (3.3) | 6,729 | 24.0 (3.6) | 2,790 | 25.6 (3.2) | 3,254 | 24.6 (4.2) |
| in 2011 | 3,628 | 26.7 (3.9) | 4,536 | 25.8 (2.9) | 1,246 | 26.9 (4.0) | 1,452 | 26.1 (4.7) |
| Leisure-time physical activity | | |  |  |  |  |  |  |
| Metabolic equivalent (MET) index (MET h/day) | | | |  |  |  |  |  |
| in 1975 | 10,309 | 2.7 (3.6) | 10,636 | 2.1 (2.5) | 3,900 | 2.6 (3.3) | 4,184 | 2.0 (2.2) |
| in 1981 | 9,780 | 2.8 (3.7) | 10,366 | 2.3 (2.5) | 3,766 | 2.8 (3.6) | 4,104 | 2.3 (2.3) |
| in 1990 | 5,620 | 3.2 (3.6) | 6,690 | 3.3 (3.2) | 2,786 | 3.1 (3.4) | 3,225 | 3.1 (3.0) |
| in 2011 | 3,374 | 2.9 (2.9) | 4,215 | 3.2 (2.7) | 1,178 | 2.6 (2.7) | 1,349 | 3.0 (2.6) |
| SD, standard deviation | | | | | | | | |

**Supplementary Table 3.** Genetic and unique environmental correlations between body mass index and leisure-time physical activity estimated using bivariate Cholesky twin model.

|  | *r* (95% CI) ^a,b^ | Number of  MZ / DZ pairs | *r_A_* (95% CI) | *r_E_* (95% CI) |
| --- | --- | --- | --- | --- |
| All |  |  |  |  |
| in 1975 | -0.05 (-0.07, -0.04) | 2,750 / 5,909 | -0.06 (-0.09, -0.03) | -0.05 (-0.08, -0.02) |
| in 1981 | -0.08 (-0.09, -0.06) | 2,578 / 5,340 | -0.06 (-0.09, -0.02) | -0.10 (-0.15, -0.06) |
| in 1990 | -0.16 (-0.17, -0.14) | 1,553 / 2,937 | -0.22 (-0.28, -0.17) | -0.12 (-0.16, -0.08) |
| in 2011 | -0.26 (-0.28, -0.24) | 870 / 1,525 | -0.33 (-0.40, -0.26) | -0.16 (-0.22, -0.11) |
| Men |  |  |  |  |
| in 1975 | -0.10 (-0.12, -0.08) | 1,231 / 2,851 | -0.12 (-0.16, -0.08) | -0.06 (-0.10, -0.02) |
| in 1981 | -0.10 (-0.12, -0.08) | 1,120 / 2,511 | -0.13 (-0.18, -0.07) | -0.08 (-0.14, -0.01) |
| in 1990 | -0.16 (-0.18, -0.13) | 638 / 1,291 | -0.23 (-0.32, -0.15) | -0.11 (-0.17, -0.05) |
| in 2011 | -0.21 (-0.23, -0.18) | 343 / 621 | -0.23 (-0.34, -0.11) | -0.18 (-0.26, -0.10) |
| Women |  |  |  |  |
| in 1975 | -0.07 (-0.08, -0.05) | 1,519 / 3,058 | -0.09 (-0.13, -0.05) | -0.02 (-0.07, 0.03) |
| in 1981 | -0.11 (-0.12, -0.09) | 1,458 / 2,829 | -0.11 (-0.16, -0.05) | -0.12 (-0.17, -0.07) |
| in 1990 | -0.16 (-0.18, -0.14) | 915 / 1,646 | -0.22 (-0.29, -0.15) | -0.13 (-0.19, -0.08) |
| in 2011 | -0.28 (-0.31, -0.26) | 527 / 904 | -0.38 (-0.47, -0.29) | -0.16 (-0.23, -0.08) |
| *r*, phenotypic correlation; CI, Confidence Interval; MZ, monozygotic; DZ, dizygotic;  *r_A_*, genetic correlation; *r_E_*, unique environmental correlation.  ^a^ CIs were corrected for family relatedness.  ^b^ The correlations were estimated using full information maximum likelihood in full cohort (n = 22,696) and in men (n = 11,282) and women (n = 11,414). | | | | |

**Supplementary Table 4.** Model-fit of the bidirectional Direction of Causation models in comparison to the models with constrained causal paths.

|  |  | BIC | Chi-squared | df | *p* | SC | CFI | TLI | RMSEA | SRMR | Chi-squared difference test (df=1)^a^ | *p* |
| --- | --- | --- | --- | --- | --- | --- | --- | --- | --- | --- | --- | --- |
| **All** |  |  |  |  |  |  |  |  |  |  |  |  |
| In 1975  (8,659 pairs) | Model 1: Bidirectional | 171532 | 97.97 | 20 | <0.001 | 2.41 | 0.98 | 0.99 | 0.030 | 0.062 |  |  |
|  | *g1* = 0 (causal effect LTPA → BMI estimated) | 171526 | 102.27 | 21 | <0.001 | 2.33 | 0.98 | 0.99 | 0.030 | 0.062 | 3.00 | 0.083 |
|  | *g2* = 0 (causal effect BMI → LTPA estimated) | 171524 | 101.24 | 21 | <0.001 | 2.33 | 0.98 | 0.99 | 0.030 | 0.062 | 0.00 | 1.000 |
| In 1981  (7,918 pairs) | Model 1: Bidirectional | 159370 | 52.67 | 20 | <0.001 | 2.2 | 0.99 | 0.99 | 0.020 | 0.043 |  |  |
|  | *g1* = 0 (causal effect LTPA → BMI estimated) | 159361 | 53.97 | 21 | <0.001 | 2.16 | 0.99 | 0.99 | 0.020 | 0.043 | 0.52 | 0.473 |
|  | *g2* = 0 (causal effect BMI → LTPA estimated) | 159373 | 59.35 | 21 | <0.001 | 2.16 | 0.99 | 0.99 | 0.021 | 0.044 | **9.06** | **0.003** |
| In 1990  (4,490 pairs) | Model 1: Bidirectional | 94649 | 19.93 | 20 | 0.463 | 1.99 | 1.00 | 1.00 | 0.000 | 0.036 |  |  |
|  | *g1* = 0 (causal effect LTPA → BMI estimated) | 94663 | 32.13 | 21 | 0.057 | 1.94 | 0.99 | 1.00 | 0.015 | 0.041 | **24.12** | **< 0.001** |
|  | *g2* = 0 (causal effect BMI → LTPA estimated) | 94641 | 20.86 | 21 | 0.468 | 1.94 | 1.00 | 1.00 | 0.000 | 0.036 | 0.86 | 0.354 |
| In 2011  (2,395 pairs) | Model 1: Bidirectional | 49890 | 31.32 | 20 | 0.051 | 1.31 | 0.99 | 0.99 | 0.022 | 0.056 |  |  |
|  | *g1* = 0 (causal effect LTPA → BMI estimated) | 49908 | 51.72 | 21 | <0.001 | 1.29 | 0.97 | 0.98 | 0.035 | 0.063 | **28.87** | **< 0.001** |
|  | *g2* = 0 (causal effect BMI → LTPA estimated) | 49882 | 31.79 | 21 | 0.062 | 1.29 | 0.99 | 0.99 | 0.021 | 0.056 | 0.00 | 1.000 |
| **Men** |  |  |  |  |  |  |  |  |  |  |  |  |
| In 1975  (4,082 pairs) | Model 1: Bidirectional | 82460 | 79.08 | 20 | <0.001 | 1.99 | 0.97 | 0.98 | 0.038 | 0.082 |  |  |
|  | *g1* = 0 (causal effect LTPA → BMI estimated) | 82463 | 86.88 | 21 | <0.001 | 1.94 | 0.96 | 0.98 | 0.039 | 0.085 | **11.88** | **0.001** |
|  | *g2* = 0 (causal effect BMI → LTPA estimated) | 82452 | 81.4 | 21 | <0.001 | 1.94 | 0.97 | 0.98 | 0.038 | 0.082 | 0.582 | 0.446 |
| In 1981  (3,631 pairs) | Model 1: Bidirectional | 74384 | 37.83 | 20 | 0.0093 | 1.92 | 0.99 | 0.99 | 0.022 | 0.093 |  |  |
|  | *g1* = 0 (causal effect LTPA → BMI estimated) | 74382 | 41.34 | 21 | 0.0051 | 1.9 | 0.98 | 0.99 | 0.023 | 0.095 | **3.94** | **0.047** |
|  | *g2* = 0 (causal effect BMI → LTPA estimated) | 74376 | 38.4 | 21 | 0.0116 | 1.9 | 0.99 | 0.99 | 0.021 | 0.093 | 0.22 | 0.641 |
| In 1990  (1,929 pairs) | Model 1: Bidirectional | 39909 | 15.08 | 20 | 0.771 | 1.79 | 1.00 | 1.01 | 0.000 | 0.055 |  |  |
|  | *g1* = 0 (causal effect LTPA → BMI estimated) | 39911 | 21.00 | 21 | 0.459 | 1.75 | 1.00 | 1.00 | 0.000 | 0.056 | **10.27** | **0.001** |
|  | *g2* = 0 (causal effect BMI → LTPA estimated) | 39902 | 15.46 | 21 | 0.799 | 1.75 | 1.00 | 1.01 | 0.000 | 0.055 | 0.07 | 0.799 |
| In 2011  (964 pairs) | Model 1: Bidirectional | 20155 | 18.64 | 20 | 0.545 | 1.32 | 1.00 | 1.00 | 0.000 | 0.078 |  |  |
|  | *g1* = 0 (causal effect LTPA → BMI estimated) | 20150 | 20.52 | 21 | 0.488 | 1.3 | 1.00 | 1.00 | 0.000 | 0.077 | 2.30 | 0.129 |
|  | *g2* = 0 (causal effect BMI → LTPA estimated) | 20148 | 19.26 | 21 | 0.568 | 1.3 | 1.00 | 1.00 | 0.000 | 0.078 | 0.48 | 0.488 |
| **Women** | |  |  |  |  |  |  |  |  |  |  |  |
| In 1975  (4,577 pairs) | Model 1: Bidirectional | 86974 | 41.73 | 20 | 0.003 | 2.85 | 0.99 | 0.99 | 0.022 | 0.073 |  |  |
|  | *g1* = 0 (causal effect LTPA → BMI estimated) | 86973 | 45.51 | 21 | 0.002 | 2.76 | 0.99 | 0.99 | 0.023 | 0.074 | **7.00** | **0.008** |
|  | *g2* = 0 (causal effect BMI → LTPA estimated) | 86967 | 43.22 | 21 | 0.003 | 2.77 | 0.99 | 0.99 | 0.022 | 0.073 | 0.67 | 0.412 |
| In 1981  (4,287 pairs) | Model 1: Bidirectional | 82829 | 19.66 | 20 | 0.479 | 2.09 | 1.00 | 1.00 | 0.000 | 0.047 |  |  |
|  | *g1* = 0 (causal effect LTPA → BMI estimated) | 82824 | 21.69 | 21 | 0.418 | 2.04 | 1.00 | 1.00 | 0.004 | 0.047 | 3.04 | 0.081 |
|  | *g2* = 0 (causal effect BMI → LTPA estimated) | 82827 | 23.04 | 21 | 0.342 | 2.05 | 1.00 | 1.00 | 0.007 | 0.048 | **4.91** | **0.027** |
| In 1990  (2,561 pairs) | Model 1: Bidirectional | 54342 | 20.13 | 20 | 0.450 | 2.11 | 1.00 | 1.00 | 0.002 | 0.048 |  |  |
|  | *g1* = 0 (causal effect LTPA → BMI estimated) | 54346 | 26.28 | 21 | 0.196 | 2.05 | 0.99 | 1.00 | 0.014 | 0.051 | **13.41** | **< 0.001** |
|  | *g2* = 0 (causal effect BMI → LTPA estimated) | 54335 | 21.23 | 21 | 0.445 | 2.05 | 1.00 | 1.00 | 0.003 | 0.048 | 1.23 | 0.267 |
| In 2011  (1,431 pairs) | Model 1: Bidirectional | 29648 | 38.23 | 20 | 0.008 | 1.28 | 0.97 | 0.98 | 0.036 | 0.079 |  |  |
|  | *g1* = 0 (causal effect LTPA → BMI estimated) | 29667 | 58.89 | 21 | <0.001 | 1.28 | 0.94 | 0.97 | 0.050 | 0.090 | **20.66** | **< 0.001** |
|  | *g2* = 0 (causal effect BMI → LTPA estimated) | 29641 | 38.78 | 21 | 0.0104 | 1.27 | 0.97 | 0.98 | 0.034 | 0.079 | 0.30 | 0.586 |
| BIC, Bayesian information criterion; df, degrees-of-freedom; SC, scaling correction; CFI, the comparative fit index; TLI, the Tucker–Lewis index; RMSEA, the root mean square error of approximation; SRMR, the standardised root-mean-square residual.  *g1* = 0: Causal path from BMI to LTPA was set to zero.  *g2* = 0: Causal path from LTPA to BMI was set to zero.  ^a^ Model fit was compared to the model-fit of the bidirectional model. | | | | | | | | | | | | |
